# Genomics reveals eleven obesity endotypes with distinct biological and phenotypic signatures

**DOI:** 10.1101/2025.06.30.25330607

**Authors:** Min Seo Kim, Minku Song, Hoyoung Kim, Sanghyeon Park, Injeong Shim, Soohyun Lim, Beomsu Kim, Xingyu Chen, Yang Sui, So Mi Jemma Cho, Satoshi Koyama, Jae-Seung Yun, Pradeep Natarajan, Patrick T. Ellinor, Akl C. Fahed, Hong-Hee Won

## Abstract

Obesity, a leading global risk factor for cardiometabolic conditions, arises from multifaceted and biologically complex mechanisms^1,2^. To elucidate the full-dimensional genetic architecture underlying obesity, we conducted a multi-trait, multi-ancestry, genome-wide association study (GWAS) by combining genetic data on anthropometric traits (body mass index, waist circumference, waist-to-hip ratio, and hip circumference) from >2 million ancestrally diverse participants. We identified 743 significant loci, including 86 previously unreported loci, representing a 13% increase in locus discovery. We leveraged machine learning and multimodal data integration to identify the likely effector genes at obesity-associated loci. We performed genetic clustering on biologically enriched multi-trait GWAS data based on Bayesian non-negative matrix factorisation^3^ in the UK Biobank (*n* = 408,816) and identified 11 obesity clusters (endotypes). In addition to recapitulating the endotypes that drive classical metabolically healthy and unhealthy obesity phenotypes, we identified nine additional obesity endotypes driven by insulin physiology, beta cell compensation, immune dysregulation, neuroendocrine regulation, and lipid metabolism. Each cluster not only was characterised by unique biomarker features and clinical trajectories but also showed cluster-specific enrichment in tissue and single-cell regulatory regions. To facilitate the clinical and research adoption of obesity endotyping, we created partitioned polygenic scores for the 11 obesity endotypes, which we externally validated their performance using the Mass General Brigham Biobank (*n* = 48,377), and made publicly available in the PGS Catalog. This study marks a step forward in both the biological resolution and clinical translation of heterogeneity in obesity, implicating that obesity prevention and management should be as diverse as the condition itself.

## Introduction

Over the past 30 years, the prevalence of obesity in children and adults has increased multiple-fold resulting in a massive global burden of disease^4,5^. With obesity reaching global epidemic proportions, decoding its biology is more important than ever, and genetic discovery has become instrumental in translating population risks into a mechanistic understanding and therapeutic development^1,6^. The landscape of gene discovery for obesity evolved dramatically with the introduction of genome-wide association studies (GWAS)^1^. In 2007, this approach yielded its first major breakthrough in obesity research, uncovering a cluster of common variants within the first intron of the *FTO* gene that showed a strong and reproducible association with body mass index (BMI)^7^. Many more GWAS followed which identified hundreds of independent loci associated with BMI.

GWAS efforts have exhaustively focused on BMI as a proxy for obesity, largely because of its convenience of measurement and widespread availability^1,8–10^. However, BMI often offers a limited lens to obesity, conflating adiposity with lean mass and obscuring the deeper biological and etiological complexity that defines the condition^1^. Since 2019, GWAS has been actively extending to diverse and more specific obesity-related traits, such as waist circumference (WC), waist-to-hip ratio (WHR), body fat percentage, fat-free mass, and imaging-derived adipose tissue^1,11,12^. While these studies typically involve smaller sample sizes compared to BMI-focused GWAS, the phenotypes often offer greater precision in capturing body-weight regulation and lead to the identification of biologically relevant loci^1^. Although each obesity trait captures distinct clinical and biological information about obesity, there has not been an attempt to combine multiple measures of obesity in discovery GWAS, which could augment power and reveal the full dimensional genetic architecture of obesity.

We conducted a multi-trait, multi-ancestry GWAS on obesity, leveraging genetic data from six ancestry groups and over 2 million participants. Genetic discovery was followed by functional annotation and fine mapping to prioritise effector genes and translate the identified loci into new biological insights. Based on this biologically enriched GWAS, we further performed genetic clustering to partition the heterogeneous aetiologies and underlying mechanisms of obesity, aiming to better characterise it as a multifaceted condition and to inform precise diagnosis and personalised care.

## Results

### Discovery of obesity loci

To investigate the complex genetic architecture underlying obesity, we performed a multi-trait GWAS using genomic structural equation modelling (SEM)^13^. Summary statistics from the GWAS of the four anthropometric traits—BMI, WC, hip circumference (HC), and WHR—were first harmonised and analysed within a hierarchical factor model for each ancestry: European (EUR), East Asian (EAS), African (AFR), Admixed American (AMR), South Asian (SAS), and Middle Eastern (MID). The genetic correlations among the four traits were generally high in non-European populations, but moderate in European and East Asian populations, demonstrating that each trait contributed distinct signals (Extended Data Fig. 1). We then conducted a multi-ancestry meta-analysis using data from ancestry-specific multi-trait GWASs to develop a comprehensive obesity GWAS (Fig. 1). This GWAS incorporated the genetic information of over 2 million participants from 15 cohorts, including GIANT, UK Biobank, FinnGen, and the Million Veterans Program, representing the largest genetic effort for obesity to date (Supplementary Table 1). We identified 743 genome-wide significant loci, including 86 previously unreported loci, representing a 13% increase in locus discovery.

**Fig. 1.**
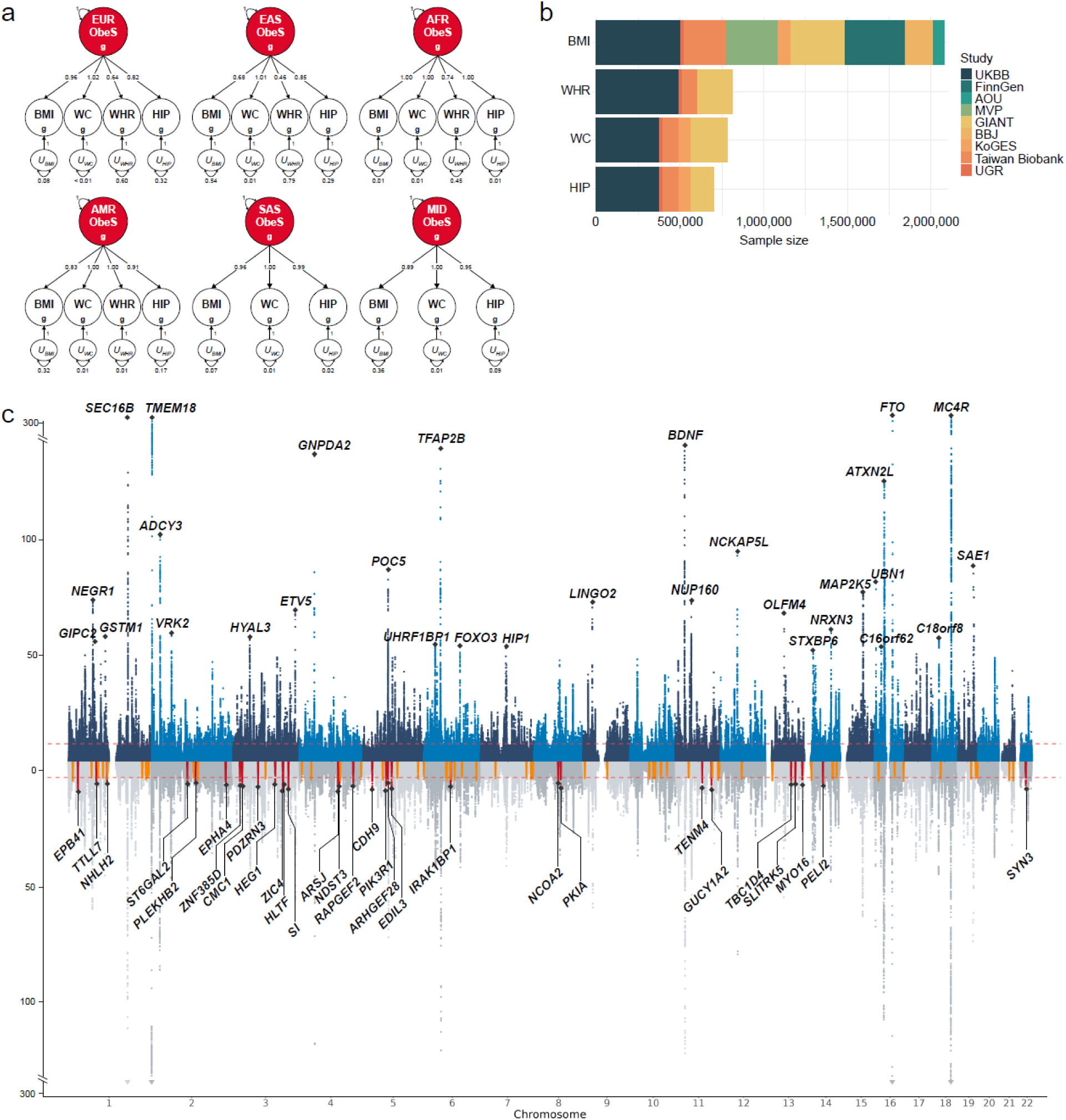
Multivariate common genetic factor model and multivariate GWAS meta-analysis results across obesity phenotypes. **a** The path diagram presents the standardised factor loadings in the hierarchical model, estimated using genomic SEM. U represents the residual variance that is not explained by the latent factors. Subscript g indicates that the model was built based on genetic covariances across the four anthropometric traits. This diagram includes the results for the EUR, EAS, AFR, SAS, AMR, and MID populations. **b** Number of GWAS included in the analysis by study and trait. **c**, Mirrored Manhattan plot of obesity showing −log_10_(*P*) for genetic associations from genomic SEM-based GWAS (y-axis) across genetic variants ordered by chromosome and base-pair positions (x-axis). In the upper mirrored plot (previously reported loci), the top 30 loci were annotated with gene symbols based on the FLAMES results. When the FLAMES results were unavailable for a given locus, the gene with the highest V2G score was selected for annotation. In the lower mirrored plot (previously unreported loci), red peak indicates the top 30 unreported loci and orange peak indicates all remaining previously unreported loci.

Tissue-specific enrichment analysis across multiple tissues based on gene expression using stratified linkage disequilibrium score regression demonstrated the enrichment of significant loci in specific brain regions (Extended Data Fig. 2), replicating previous findings^8^. Our finding confirmed the attribution of brain or central nervous system as a principal driver of obesity by controlling energy intake and expenditure^1,14,15^. We used machine learning and multimodal data integration to identify likely effector genes at obesity-associated loci^16^ (Supplementary Table 3). Strong signals in previously unreported loci include *EPB41*, *EPHA4*, *CMC1*, *ZIC4*, *HLTF*, *ARSJ*, *NDST3*, *PIK3R1*, *PKIA*, *TENM4*, *GUCY1A2*, *MYO16*, and *SYN3* (Fig. 1c)—among them are genes known to be associated with neural development, synaptic connectivity, immune regulation, and neuroendocrine controls^17–19^.

### Non-European specific loci

While the multi-ancestry GWAS served as a groundwork for downstream analysis, such as genetic clustering, we also scrutinised non-European specific genetic loci to identify potential new drug targets. Drug targets can be discovered from genetic studies of non-Europeans–for instance, *PCSK9* (target for PCSK9 inhibitor) from Lebanese^20^–due to the high minor allele frequency of functional variants of the corresponding gene in certain ancestry groups. To evaluate ancestry-specific signals at obesity-associated loci, we performed cross-population comparisons based on lead variants defined through multi-ancestral GWAS meta-analysis using genomic SEM. The set of loci used for comparison was established from this unified framework, enabling the consistent identification of overlapping and population-specific signals across the EUR, EAS, and AFR populations. Other ancestry groups, including the MID, SAS, and AMR populations, were excluded from the cross-population comparison because of the lack of significant genome-wide associations in their respective ancestry-specific GWAS results.

As illustrated in Extended Data Fig. 3, the majority of loci were ancestry-specific and predominantly European-specific, likely due to the larger GWAS sample size. In the pairwise comparison between EUR and EAS, 532 loci were identified as EUR-specific, 9 were unique to EAS, and 127 loci were shared between the two populations. When comparing the EUR and AFR, we observed 644 EUR-specific loci, 14 AFR-specific loci, and only one shared locus. In the combined analysis across all three populations, 528 loci were exclusive to EUR, nine to EAS, and one to AFR (Supplementary Table 8). Although the allele frequencies of ancestry-specific loci varied, their effect sizes were similar across ancestries (correlation of effect sizes: *R*^2^ = 0.907 for EUR vs. EAS and *R*^2^= 0.890 for EUR vs. AFR). These results confirm the value of incorporating diverse ancestry groups into genetic studies to uncover signals that may remain undetected in EUR-centric analyses and to enhance the generalisability of genetic discoveries.

### Obesity clusters

A multi-trait, multi-ancestry approach yielded 11 obesity genetic clusters (endotypes). We identified these clusters using Bayesian non-negative matrix factorisation (bNMF) applied to multi-trait GWAS summary statistics, which grouped variants with shared phenotypic signatures. The phenotypic features and biomarker profiles of each cluster were generally consistent between sexes (Extended Data Figs. 4 and 5). As this is the first study to disclose endotypes for obesity at this scale, we put forth extensive descriptions for each cluster based on the current understanding of the underlying genes and mechanisms.

### Metabolically unhealthy cluster

This cluster exhibits a constellation of increased adiposity across multiple compartments: abdominal subcutaneous adipose tissue (ASAT+), visceral adipose tissue (VAT+), gluteofemoral adipose tissue (GFAT+), liver fat accumulation (Liver fat+), and trunk fat percentage (TFP+). This pattern represents systemic adiposity with a concerning distribution in metabolically harmful depots, particularly in visceral and ectopic (liver) fat. It likely represents the classic “metabolically unhealthy obesity (MUO)” phenotype, which is evident by umbrella increments of risk for multiple comorbidities in Fig. 2. The GWAS-partitioned polygenic score (pPS) for the cluster was associated with increased levels of all MRI traits, including epicardial adipose tissue, ASAT, VAT, GFAT, liver fat percentage, and pancreas fat percentage (Fig. 3). The primary pathophysiology may involve dysregulation of the hypothalamic control of food intake and energy expenditure (*FTO* and *BDNF* cluster-enriched genes; Fig. 2 and Table 1)^21,22^, leading to a positive energy balance and adipose tissue expansion. Consistent with this, our cluster-specific pathway analysis revealed significant enrichment for nervous system development and brain glutamatergic synapse pathways (Supplementary Table 25), as well as the highest enrichment of open chromatin regions in glutamatergic neurons (Supplementary Table 21). The expanded VAT can become inflamed, releasing pro-inflammatory cytokines and excess free fatty acids directly into portal circulation, causing hepatic insulin resistance and steatosis^23^. Mitochondrial dysfunction (*TOMM40*) may compound this by reducing fatty acid oxidative capacity and promoting further ectopic fat deposition^24^. The epigenetic regulators (*H3C8* and *H2AC6*) suggest that environmental factors may interact with genetic predisposition, potentially explaining how lifestyle factors exacerbate genetic risk in metabolically unhealthy obesity^25,26^; Wahl et al. demonstrated that DNA methylation alterations are the consequence of fat accumulation, rather than the cause^25^.

**Fig. 2.**
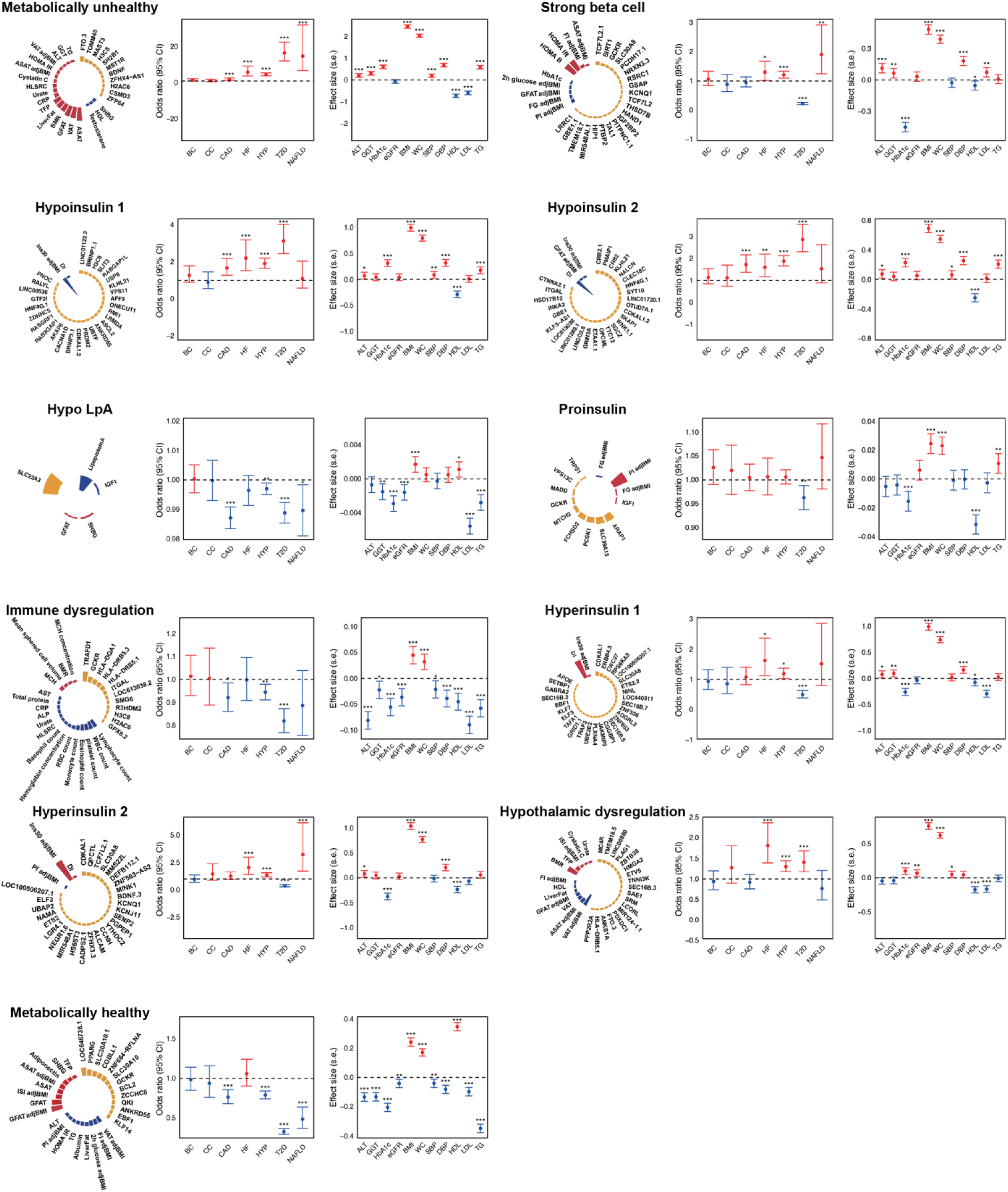
Characteristics of multi-ancestry obesity genetic clusters and their associations with continuous traits and clinical outcomes. Each plot illustrates the top-weighted loci and traits within each multi-ancestry obesity genetic cluster along with cluster associations with selected continuous traits and clinical outcomes in UK Biobank. In the circular plot, the bar length represents the cluster weight, as determined by the bNMF algorithm. The yellow bars indicate genetic loci, red bars represent traits with positive beta, and blue bars represent traits with negative beta within each cluster. A maximum of 30 elements (loci and traits) with the highest weights are displayed per cluster while all others are provided in the Supplementary Materials. The centre panel presents the associations of partitioned polygenic score (pPS) with clinical outcomes, where each dot represents the odds ratio (ORs) per 1 standard deviation (s.d.) increase. Error bars denote 95% confidence interval (CIs). The right panel of each cluster plot depicts the association between individual-level pPS and continuous traits. Error bars indicate the standard error (s.e.) from a linear regression model. Across all components, positive associations are shown in red, and negative associations are shown in blue. The presented *P* values were derived from two-sided association tests, as described in the **Methods** section. In the Proinsulin cluster, FG.adjBMI(+) was associated with rs11606287, and FG.adjBMI(−) was associated with rs7123876, whose effect sizes were similar but acted in opposite directions.

**Fig. 3.**
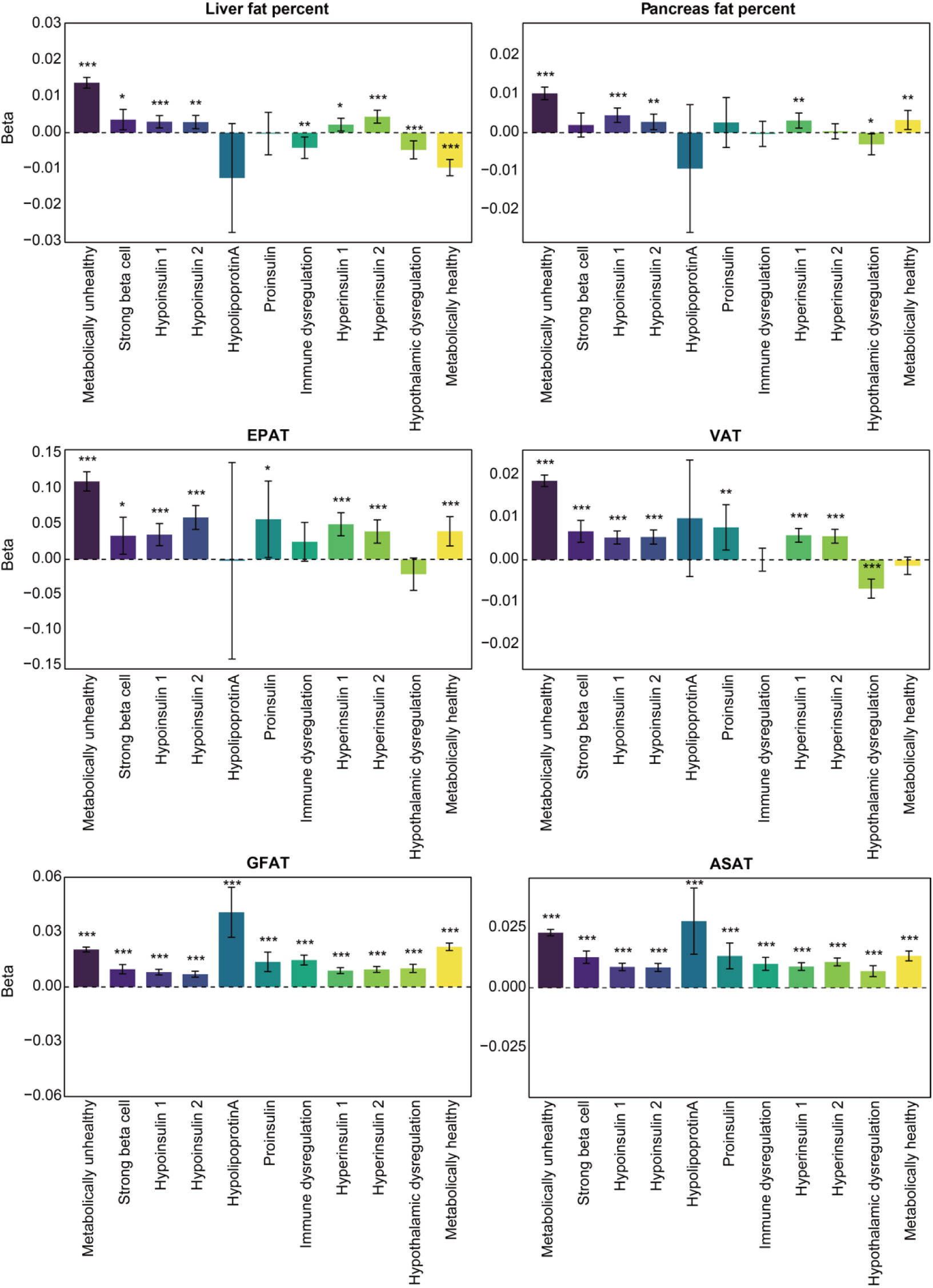
Exploring the associations between multi-ancestry obesity genetic clusters and obesity-related imaging traits. Each plot illustrates the association between GWAS-partitioned polygenic scores (pPS) and obesity-related imaging traits. Each bar represents the effect size (z-score) of a genetic cluster on a given trait, with clusters shown on the x-axis. The y-axis indicates the effect size (standard error, s.e.), where positive values indicate a stronger association in the positive direction, and negative values indicate the opposite. Error bars represent 95% confidence intervals. The colour of each bar corresponds to a specific genetic cluster. Asterisks above the bars denote statistical significance (\**P* < 0.05, \*\**P* < 0.01, \*\*\**P* < 0.001). The *P* values were derived from two-sided association tests. EPAT, epicardial adipose tissue; ASAT, abdominal subcutaneous adipose tissue; VAT, visceral adipose tissue; GFAT, gluteofemoral adipose tissue.

**Table 1.**
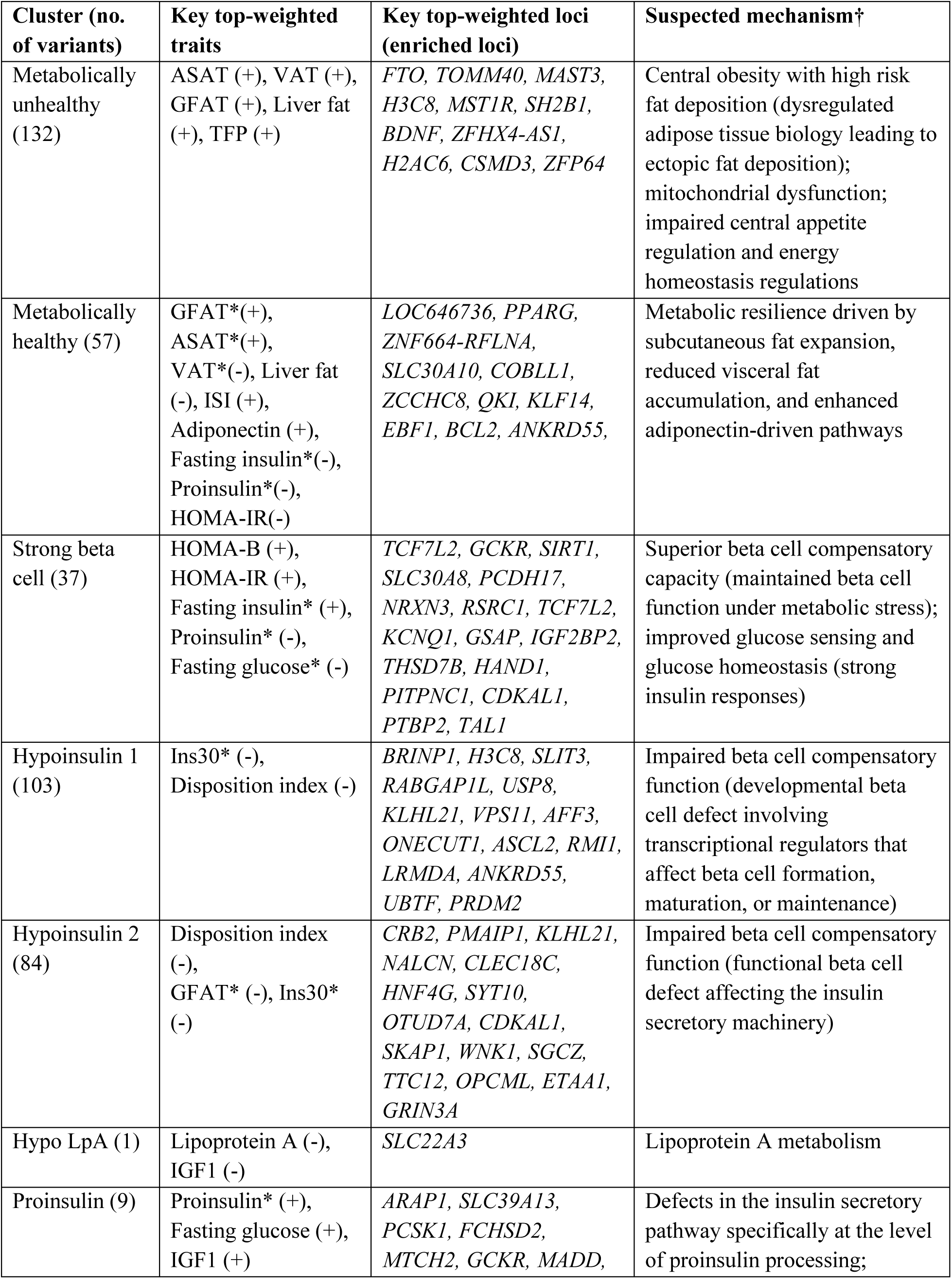
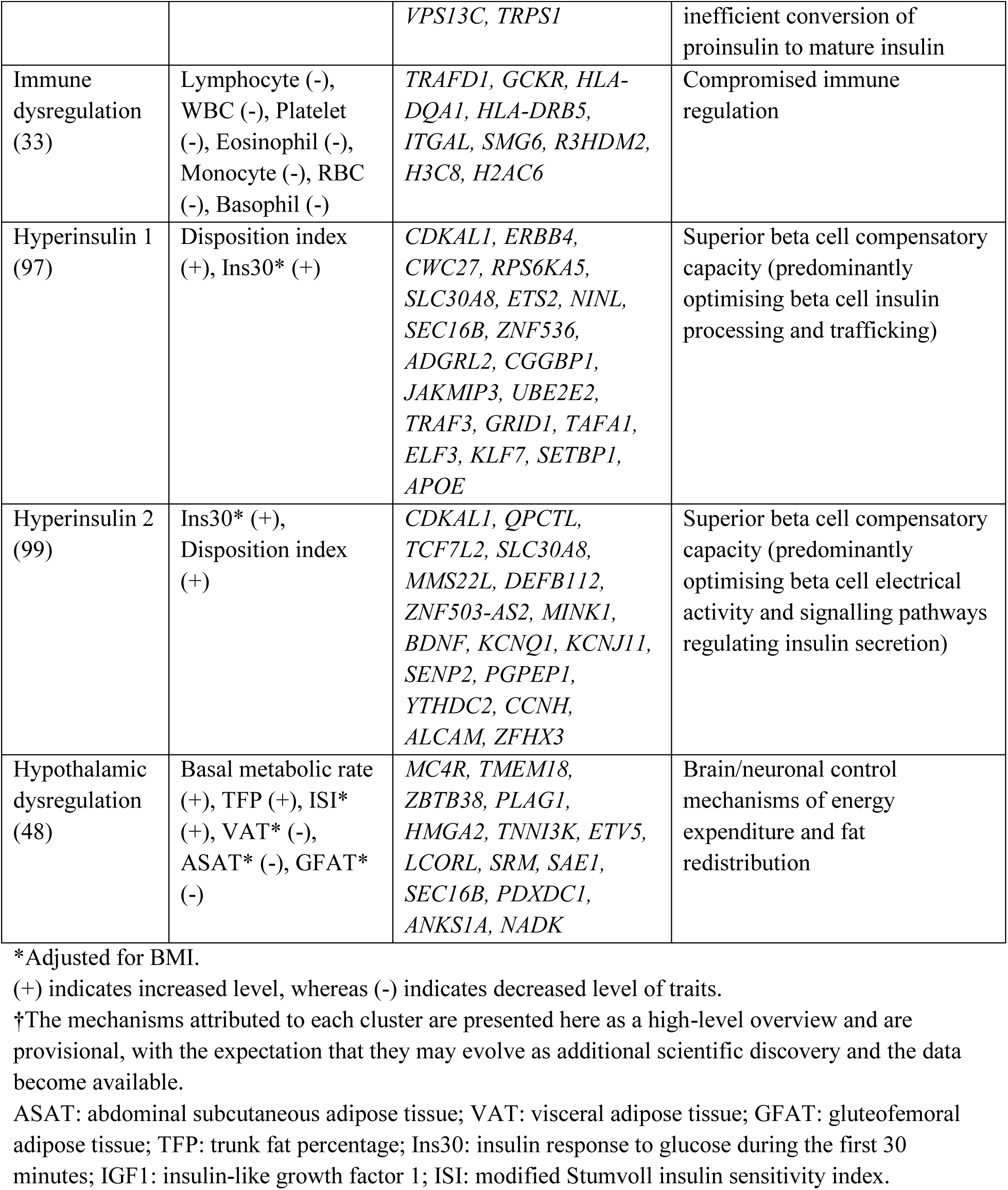
Overview of multi-ancestry obesity genetic clusters.

### Strong beta cell cluster

This cluster represents a remarkable compensatory phenotype characterised by HOMA-B (+), HOMA-IR (+), fasting insulin (+), proinsulin (−), and fasting glucose (−). This cluster likely represents individuals with a genetically determined superior beta cell adaptation capacity, allowing them to maintain normal glucose levels despite significant insulin resistance (HOMA-IR+). The genetic profile supports a “super-compensator” phenotype through enhanced beta cell proliferation and survival (*TCF7L2* and *SIRT1*)^27–29^, optimised glucose sensing (*GCKR*)^30^, and efficient insulin processing and secretion (*SLC30A8*, *CDKAL1*, *IGF2BP2*)^31,32^. The low proinsulin-to-insulin ratio despite increased insulin production suggests highly efficient proinsulin processing machinery, likely through enhanced *CDKAL1* function improving translational fidelity and *SLC30A8* optimising insulin packaging^32^. This aligns with our finding that this cluster was associated with a reduced risk of type 2 diabetes (T2D) and low HbA1c levels, despite positive associations with BMI and WC (Fig. 2). The lower risk of T2D in this cluster was validated using an external dataset (MGB Biobank). High enrichment of open chromatin regions was demonstrated in fetal endocrine cell, fetal islet, adult beta 1, delta, gamma, and adult enterochromaffin cell, to mention a few (Fig. 4 and Supplementary Tables 20–21)

**Fig. 4.**
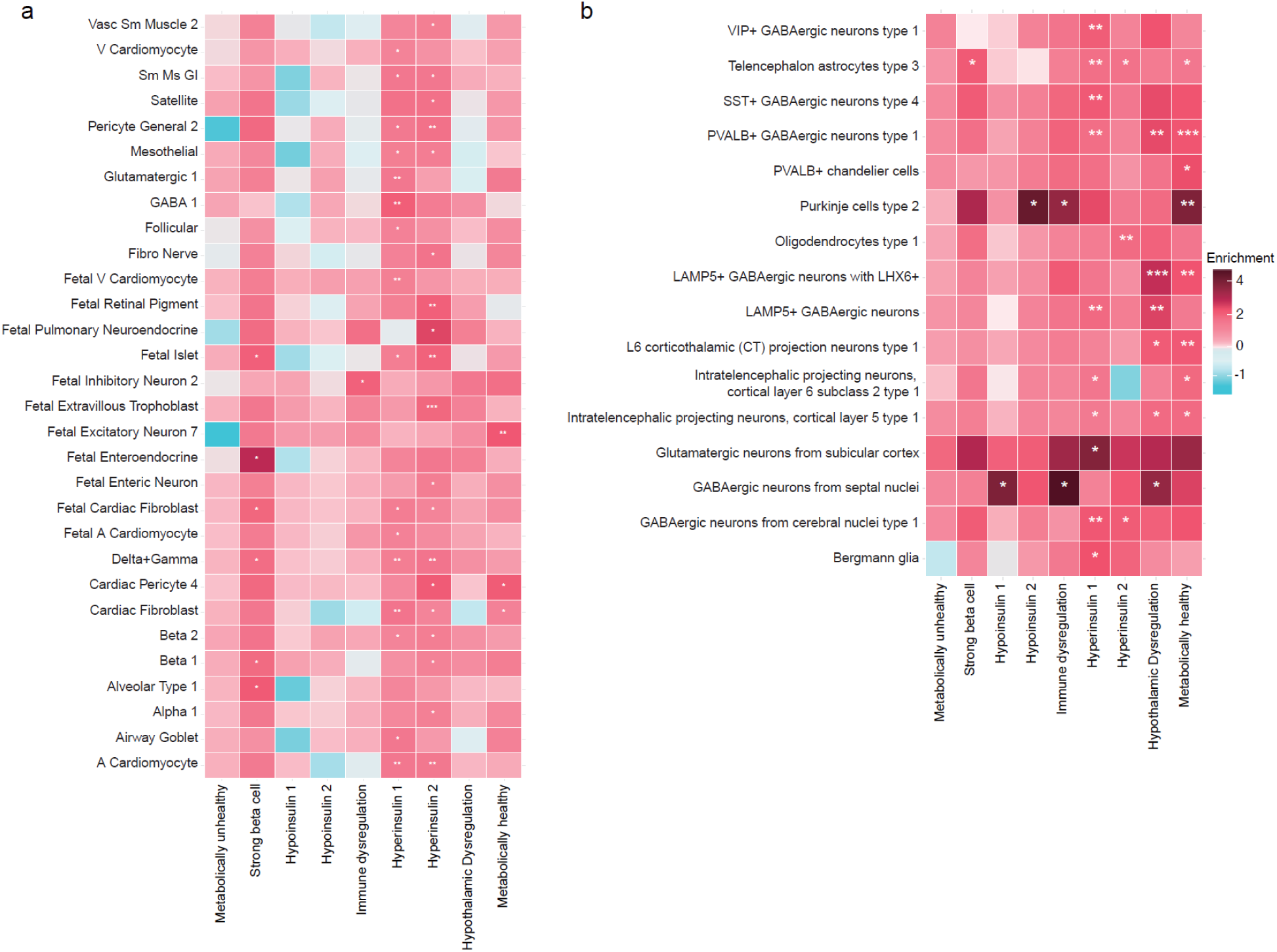
Heat map of cluster-specific enrichments of obesity associations for cell-type-specific regions of open chromatin derived from single-cell ATAC-seq peaks in adult and fetal tissues and brain tissue. **a**, **b** Heatmaps show significant cluster-specific enrichment of genomic annotations in single-cell accessible chromatin data from 222 cell types across 30 human adult tissues and 15 human fetal tissues (**a**) and from 106 cell types in the human brain (**b**). Columns represent obesity genetic clusters, whereas rows correspond to cell types that were significantly enriched for obesity associations in at least one cluster. *q*-values were corrected for FDR, with significance thresholds indicated as *FDR < 0.05, **FDR < 0.01, and ***FDR < 0.001. Only cell types exhibiting significant signals (FDR < 0.05) in at least one cluster are shown here, whereas the complete results for all cell types and clusters are provided in the Supplementary Materials.

### Hypoinsulin 1 and 2 clusters

These clusters are characterised by impaired early insulin secretion and a negative disposition index, representing a primary beta cell dysfunction phenotype (Fig. 1 and Table 1). The central pathophysiological mechanism of Hypoinsulin 1 appears to involve the transcriptional dysregulation of beta cell development, differentiation, and maturation, with *ONECUT1* potentially playing a pivotal role^33^. In contrast, Hypoinsulin 2 may represent a functional beta cell defect affecting the stimulus-secretion coupling pathway rather than beta cell development or differentiation. The enrichment of genes encoding multiple ion channels and calcium-handling proteins (*NALCN*, *GRIN3A*, *WNK1*, *SYT10*) suggests dysregulation of electrical and calcium signalling essential for glucose-stimulated insulin secretion^34^. The negative GFAT phenotype in Hypoinsulin 2 suggests altered adipose tissue distribution, possibly due to subclinical insulin deficiency affecting adipose development patterns. Unlike Hypoinsulin 1, which appears to involve a developmental defect in beta cell formation, Hypoinsulin 2 likely represents individuals with normal or suboptimal beta cell development, but with more prominent dysfunctional glucose sensing and insulin secretory machinery. These clusters were enriched in nervous system development and neuronal differentiation (Supplementary Table 25), possibly indicating an underlying neuroendocrine axis. Both clusters were associated with an increased risk of T2D (Fig. 2), and this finding was consistent with the external validation (Fig. 5). These clusters were also associated with earlier onset of obesity (Fig. 5).

**Fig. 5.**
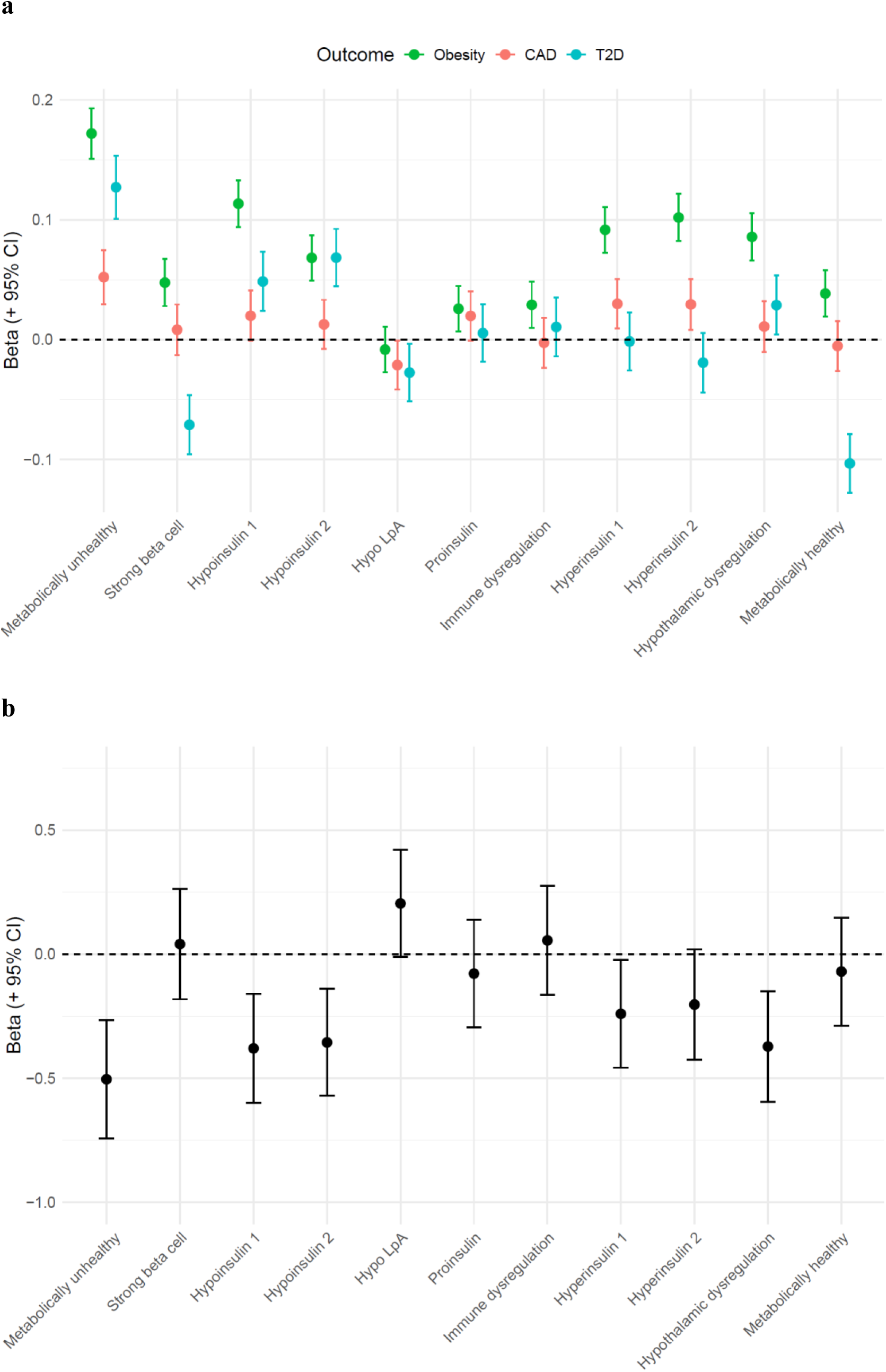
Association of partitioned polygenic scores (pPS) with metabolic outcomes and age of obesity onset in 48,377 individuals in the MGB Biobank. **a** Association between 11 obesity pPSs and metabolic outcomes, presented as beta per s.d. of pPS, estimated from a multivariate logistic regression model adjusted for age, sex, and ancestry. **b,** Association between 11 obesity pPSs and age at diagnosis of obesity, presented as beta (in years) per s.d. of pPS, estimated from a multivariable linear regression model adjusted for sex and ance

### Hypo LpA cluster

This cluster exhibited decreased level of lipoprotein A (Lp(a)) and is defined by a single gene, *SLC22A3*. The *SLC22A3* and *LPA* are genomically adjacent and known to share regulatory mechanisms^35^. Genetic variants of *SLC22A3* modulate *LPA* expression, possibly lowering the level of Lp(a)^35,36^. This could partially explain the decreased risks of coronary artery disease (CAD), T2D, and many other traits in this cluster (Fig. 2). Importantly, clusters are dynamic rather than static, as evidenced by T2D genetic clustering, in which some clusters have merged or split in recent updates as larger multi-ancestry GWAS become available^3,37^. Interestingly, the LpA cluster with single variants was also discovered in an early T2D clustering effort by Kim et al.^37^, but later merged with the SHBG cluster to form the SHBG-LpA cluster^3^.

### Proinsulin cluster

This cluster displayed elevated proinsulin (+), fasting glucose (+), and IGF1 (+) levels, likely representing a specific defect in insulin processing rather than in overall beta cell function (Fig. 1 and Table 1). *PCSK1* deficiency is likely the primary driver as it directly impairs prohormone processing^38^. The disproportionate elevation in proinsulin levels might suggest inefficient proinsulin-to-insulin conversion, creating a relative insulin deficiency despite adequate insulin gene transcription and translation. This is compounded by defects in vesicle trafficking (*MADD* and *VPS13C*) and reduced glucose-stimulated insulin granule docking and exocytosis (*SLC39A13/ZIP3*), which create a “traffic jam” in the insulin secretory pathway^39,40^. Insulin regulates appetite via brain signalling, whereas proinsulin lacks this function^41^. When proinsulin is not efficiently converted into insulin, the absence of insulin satiety signals can result in increased food intake and weight gain^41^. The gene set for Proinsulin cluster was most prominently expressed in macrophages in adipose and pancreatic tissues (Extended Data Fig. 6 and Supplementary Table 26). While the Proinsulin cluster was associated with reduced T2D risk in the UK Biobank (Fig. 2), this finding was not validated in the MGB Biobank (Fig. 5). It may be prudent to retain the null hypothesis until further data suggest otherwise.

### Immune dysregulation cluster

The cluster represents a profound disturbance in immune-metabolic crosstalk, characterised by a global reduction across multiple immune cell lineages (Fig. 1 and Table 1). In a healthy state, type 2 immune responses (involving eosinophils, macrophages, and innate lymphoid cells) predominate in lean adipose tissue and actively promote metabolic homeostasis^42^. The global reduction in eosinophils and lymphocytes observed in this cluster may reflect a compromised type 2 immune response, disrupting the finely tuned immune milieu necessary for adipose tissue homeostasis and metabolic resilience^42^ and facilitating obesity development. Genetic profiling revealed *TRAFD1* is a pivotal regulator in this cluster (Table 1). This gene is known to act as a trans-regulator of 41 genes involved in IFNγ signalling and MHC class I antigen processing/presentation pathways, suggesting its potential role as a master regulator of immune activation pathways^43^. High enrichment of cluster signals in open chromatin regions was detected in fetal inhibitory neuron 2 and in adult GABAergic neurons in multiple brain regions (Fig. 4 and Supplementary Tables 20–21). Although this cluster was consistently associated with an increased risk of obesity across the cohorts (Figs. 2 and 5), its associations with T2D and CAD were inconsistent.

### Hyperinsulin 1 and 2 clusters

These clusters demonstrated enhanced beta cell function with a positive disposition index and a positive early insulin response, indicating superior insulin secretory capacity (Fig. 1 and Table 1). For the Hyperinsulin 1 cluster, enriched genes included *CDKAL1* (protein translation in beta cells)^44^, *SLC30A8* (zinc transporter essential for insulin crystallisation)^45^, and *SEC16B* (ER-to-Golgi transport)^46^, which collectively enhance multiple components of the insulin secretory machinery at various levels, including biosynthesis, processing, granule formation, exocytosis, and beta cell survival. The positive early insulin response with a preserved disposition index suggests that these individuals may maintain excellent beta-cell glucose sensitivity despite potential insulin resistance. This may represent genetically determined resilience to diabetes development, as enhanced beta cell capacity provides a substantial reserve to compensate for increasing insulin demand.

While sharing some genes with Hyperinsulin 1 cluster (*CDKAL1* and *SLC30A8*), the genetic profile of the Hyperinsulin 2 cluster pointed more to beta cell electrical activity and regulatory pathways. Key genes include *TCF7L2* (WNT signalling affecting beta cell proliferation)^27^, potassium channels crucial for insulin secretion (*KCNQ1* and *KCNJ11*)^47^, and neurotrophin signalling (*BDNF*)^22^. The mechanism appears to focus on optimising beta cell electrical activity and signalling pathways that regulate insulin secretion, rather than protein processing and trafficking, as in the Hyperinsulin 1 cluster. While both Hyperinsulin 1 and 2 clusters were associated with a decreased risk of T2D in the UK Biobank, these findings were only replicated in the MGB Biobank with respect to point estimates (Figs. 2 and 5). This may reflect the smaller sample size of the MGB Biobank compared with that of the UK Biobank (48,377 vs. 408,816), which may result in wider confidence intervals and a greater propensity to null findings. Strong enrichment of the cluster signals in the open chromatin regions of fetal islets, beta 2 cells, delta cells, gamma cells, and many cell types in brain regions was observed in both Hyperinsulin 1 and 2 clusters, potentially implicating a regulatory role of pancreatic and brain cells in shaping the neuroendocrine circuits governing insulin physiology (Fig. 4 and Supplementary Tables 20–21).

### Hypothalamic dysregulation cluster

This cluster exhibited a distinct metabolic phenotype, characterised by an increased basal metabolic rate and TFP, along with reduced VAT, ASAT, and GFAT (Fig. 1 and Table 1). This cluster most likely represents a centrally driven metabolic phenotype in which the hypothalamic circuits controlling energy homeostasis and body composition are dysregulated. The MC4R pathway likely serves as the cornerstone, integrating leptin and insulin signals, to regulate energy balance and autonomic output to peripheral tissues^1^. The *TMEM18* gene is also well plays a role in the central control of appetite and body weight regulation^1,48^. Strong enrichment in open chromatin regions across multiple brain cell types and minimal enrichment in non-brain cell types was observed in this cluster (Fig. 4 and Supplementary Tables 20–21), recapitulating the central mechanism of this cluster. The increased basal metabolic rate coupled with the unique fat distribution pattern (increased TFP despite generally decreased fat depots) suggests enhanced sympathetic nervous system activity driving lipolysis in peripheral adipose tissue^49^. Chronic sympathetic overactivity has been suggested to drive obesity by causing beta-adrenoceptor desensitisation, reducing thermogenic capacity, and promoting insulin resistance^50^. This creates a perpetual loop in which impaired energy expenditure and insulin resistance exacerbates weight gain^50^.

### Metabolically healthy cluster

This cluster exhibited a remarkably favorable metabolic profile: GFAT+, ASAT+, VAT-, Liver fat-, insulin sensitivity+, adiponectin+, fasting insulin, proinsulin, and HOMA-IR (Fig. 1 and Table 1). The umbrella risk reduction of obesity-related morbidities such as CAD, hypertension, non-alcoholic fatty liver disease (NAFLD), and T2D was also observed. This cluster likely captures the underlying endotype of the “metabolically healthy obesity (MHO)” phenotype^51^ that shows metabolically favorable fat distribution, excellent insulin sensitivity (ISI+), and metabolic resilience despite potentially expanded adipose tissue mass^51^. The PPARG-centred mechanism promotes insulin-sensitive adipocyte differentiation with optimal adipokine secretion (adiponectin+), creating a high-quality adipose tissue that efficiently buffers lipid flux and prevents lipotoxicity in non-adipose tissues^52^. This may be complemented by the trans-regulatory effects of *KLF14* on multiple adipose-specific genes that enhance insulin sensitivity^53,54^. A significant enrichment of open chromatin regions was observed in fetal excitatory neurons, cardiac pericytes/fibroblasts, and various brain cell types (i.e., PVALB+ and LAMP5+ GABAergic neurons, and Purkinje cells) (Fig. 4 and Supplementary Tables 20–21).

### Distribution of obesity genetic clusters by ancestry

We next examined how multi-ancestry obesity genetic clusters contributed to overall obesity risk across ancestral populations. For the MGB Biobank participants, we calculated partitioned polygenic scores (pPSs) using cluster-specific weights generated by the bNMF algorithm based on obesity-related traits. We observed that the distribution of obesity clusters differed across ancestries. For instance, the median pPS for the Metabolically unhealthy cluster was highest in the EUR ancestry group and lowest in the EAS ancestry group (Extended Data Fig. 7). Furthermore, within each ancestry group, we found varied proportions of genetic obesity risk attributable to each cluster (Extended Data Fig. 8). These findings demonstrate that ancestry-specific obesity risk profiles may arise from distinct configurations of the 11 endotypes, providing a partial mechanistic basis for population-level differences in obesity-related comorbidity patterns and BMI thresholds for obesity^55^. To account for ancestry-specific effects, we regressed the top 10 principal components (PCs) from the pPS for each cluster and standardised the residuals to a normal distribution (Extended Data Fig. 8). PC-adjusted pPS was used in the downstream analyses (Fig. 5), requiring ancestry adjustment to enable direct comparisons across clustered individuals.

Notably, we used the terms cluster and endotype interchangeably because both represent mechanistically defined, non-mutually exclusive disease constructs (where an individual may exhibit multiple concurrent endotypes). This stands in contrast to ‘subtype,’ which is typically defined by observable phenotypic characteristics and are often mutually exclusive categories (for example, allergic versus non-allergic).

## Discussion

To better understand the biology and etiological heterogeneity of obesity across diverse populations, we assembled a corpus of GWASs for four obesity-related anthropometric traits from six ancestry groups and conducted the first multi-trait, multi-ancestry GWAS (>2 million participants) for obesity. We identified 743 genetic loci associated with obesity, including 86 previously unknown loci. Achieving a 13% increase in loci discovery is noteworthy considering that obesity has long been regarded as a near-saturated trait in GWAS research, with prior studies cataloguing hundreds of associated loci. By leveraging our biologically enriched GWAS data, we performed genetic clustering, as described elsewhere^3^, and discovered 11 obesity clusters. Each cluster was characterised by unique biological and biomarker features (i.e., BMI, alanine transaminase (ALT), HbA1c, systolic blood pressure (SBP), and LDL) and showed significant cluster-specific enrichment in tissue expression and single-cell regulatory regions.

This multi-trait strategy has been proven to be effective in identifying previously unreported genetic loci. Recent GWAS on depression^56^ and bipolar disorder^57^ have boosted genetic signal detection by combining GWAS from multiple phenotype assessments (i.e., electronic health records, questionnaires, and self-reported diagnoses). Our findings in obesity genetics represent another successful example of this approach, with the use of diverse obesity-related traits, resulting in a marked increase in detected genetic associations. Following the construction of a multi-trait GWAS for each ancestry, we conducted a multi-ancestry meta-analysis to produce an omnibus obesity GWAS that unified genomic insights from multiple measures and ancestries, which unveiled previously unreported 86 loci (top signals include *SKIV2L*, *MINDY3*, *PIK3R1*, *ZIC4*, *MECR*, *ARSJ*, *CCHCR1*, *GUCY1A2*, and others; Supplementary Tables 2 and 3). The relevance of these genes in obesity and metabolic regulation is yet to be thoroughly characterised, highlighting the importance of further follow-up studies to clarify their roles. Notably, multi-tissue LDSC analysis demonstrated predominant enrichment of obesity-associated loci in brain regions, consistent with a previous work^8^. Despite the unique biological signatures of each cluster, all clusters showed some degree of brain pathway involvement, reinforcing the fundamental role of the central regulation of hormones and energy balance in the pathophysiology of obesity.

The aetiologies and mechanisms delineated in our clusters recapitulate previous evidence and knowledge of obesity. Of the 11 clusters, six demonstrated a primary association with insulin regulation and pancreatic beta cell function. The large proportion of diabetes-related obesity endotypes reinforced the widely observed association between obesity and T2D, which frequently co-occur and are closely interconnected through a shared pathophysiology^58–60^. While diabetes-related obesity clusters consistently exhibited increases in BMI and WC, they demonstrated divergent effects on T2D risk, potentially reflecting the variability in brain–pancreas crosstalk. This diabetogenic heterogeneity echoes a previous genetic finding by Coral et al.^61^, who identified two distinct obesity genetic profiles with opposing diabetogenic effects. Our clusters also captured other nondiabetic aetiologies of obesity, including metabolic resilience and immune dysregulation. Metabolically healthy and unhealthy clusters appear to underlie the clinically recognised MHO and MUO phenotypes, accentuating the translational relevance of these clusters. The identification of immune dysregulation clusters corroborates previous evidence linking altered immune function to obesity^62,63^. Taken together, these clusters collectively capture an extensive body of obesity research, providing strong validation for our findings.

The MHO and MUO phenotypes represent some of the most interesting and extensively studied subtypes in obesity^2^. While evidence substantiates their biological and clinical relevance^64^, whether MHO represents a true status or merely a transient state on its way to metabolic dysfunction remained controversial^2^. Our findings on Metabolically healthy and Metabolically unhealthy clusters suggest that their existence as distinct entities might be genuine, although the clinical classification of MHO and MUO may be influenced by transitional dynamics^2,65^. Metabolically healthy cluster was characterised by favorable fat distribution (↑GFAT, ↓VAT, ↓liver fat), endocrine function (↑insulin sensitivity, ↑adiponectin, ↓proinsulin, ↓HOMA-IR), and cardiometabolic profiles (umbrella risk reduction of CAD, NAFLD, and T2D) (Fig. 2); in contrast, the Metabolically unhealthy cluster exhibited the opposite patterns, broadly recapitulating what has been known for MHO vs. MUO. Furthermore, distinct gene set and pathway enrichments between the two clusters pointed to differing genetic architectures. Cell-type regulatory landscapes also diverged, with Metabolically healthy cluster showing more pronounced enrichment across diverse cell types––particularly in brain regions––compared to Metabolically unhealthy cluster (Fig. 4). This may suggest potential developmental programming aspects that might predispose individuals to metabolically healthy or unhealthy obesity. Currently, the classification of individuals as MHO or MUO is largely retrospective and post hoc, inferred only after the manifestation of long-term metabolic outcomes. This delay limited the ability to intervene early and prevent comorbidities. Our genetic clustering framework, however, enables early, primordial stratification of these endotypes, theoretically from the initial clinical encounter based on personal genotype data.

A range of strategies has been employed to uncover the subtypes and endotypes of obesity. As expected, this endeavour has mainly focused on leveraging clinical features and biomarkers^64,66,67^. Coral et al. recently applied unsupervised clustering to clinical markers such as LDL and SBP for obesity subclassification^66^. Although clinical traits are routinely collected and contain rich medical information, they often fluctuate over the course of life and typically manifest in adulthood. Genomic data, a stable, single-assessment biomarker, have been largely overlooked in this clustering effort for obesity, which starkly contrasts with the T2D space, where eight^68^ to twelve^3^ distinct T2D genetic clusters have been identified. For obesity, Grant et al. provided early evidence for the genetic clustering of BMI; however, their primary aim was to introduce a new statistical framework, with BMI serving as a use case^69^. The identification of five clusters indicated potential genetic endotypes of obesity, although these clusters have not been sufficiently characterised^69^. Additionally, their analysis was based on a GWAS for BMI, not on comprehensive obesity phenotypes, and was limited to European populations. Our study delivered an unprecedented level of scale and resolution for obesity endotyping through the synergistic application of multi-ancestry genomics, systems biology, and clinical phenotyping. Unlike traditional phenotype-driven classifications, genetically anchored clusters can decode obesity risk and its endotypes even during the presymptomatic stages. This paradigm shift creates opportunities for age-agnostic prevention, including early life interventions informed by an individual’s genetic profile. With healthcare increasingly embracing individualised and preventive strategies, the early life identification of obesity endotypes through personal genetic data represents a compelling opportunity to advance precision medicine.

This study has several limitations that should be considered when interpreting the findings. First, despite the demonstrated value of including diverse ancestral populations in genetic analyses, our study was limited by the current availability of genetic data. To date, GWAS remain heavily weighted toward individuals of European descent, and this bias is reflected in our own biobank analyses, in which the majority of participants also had European genetic backgrounds. Consequently, the generalisability of our findings to non-European populations remains restricted. However, validation in the ancestrally diverse MGB Biobank provides some degree of reassurance and support for the robustness of our clusters across different backgrounds. Second, while we performed ancestry-stratified GWAS to identify obesity-associated loci unique to non-European populations, only a small subset of ancestry-specific loci reached genome-wide significance, likely because of the disproportionate underrepresentation of non-European individuals in current genetic studies. This suggests that critical variants with large effect sizes or high allele frequencies in non-European populations may remain undetected, potentially obscuring ancestry-specific biological pathways. Finally, the mechanistic attributes for each cluster are based on current knowledge and provisions. As the field advances and more functional and experimental data become available, mechanistic interpretations can be refined or revised.

In summary, a multi-trait, multi-ancestry GWAS achieved a 13% increase in locus discovery in obesity. Our genetic clustering based on the largest biologically enriched obesity GWAS confirmed the heterogeneity of obesity, with this biological heterogeneity directly contributing to divergent clinical phenotypes. This implies that obesity prevention and management should be as diverse as the condition itself, moving beyond uniform strategies and a one-size-fits-all approach. To ensure that obesity clusters could be widely used in clinical practice and research, we created pPS weight files for the 11 clusters available in the PGS Catalog. Obesity endotyping using genotype data could be a critical first step toward precise obesity management.

## Methods

### Selection of GWAS on obesity-related traits and meta-analysis

We focused on four obesity-related anthropometric traits (body mass index [BMI], waist circumference [WC], hip circumference [HIP], and waist-to-hip ratio [WHR]) for downstream analysis. To obtain well-powered and ancestrally diverse association statistics, the largest available GWAS summary statistics for each trait were collected from multiple biobanks and consortia. For European ancestry (EUR), we utilised data from the Genetic Investigation of Anthropometric Traits (GIANT), UK Biobank (UKBB), FinnGen (Release 12), Million Veterans Program (MVP), and All of the Us (AoU) Research Program. For East Asian ancestry (EAS), summary statistics from the Taiwan Biobank, Korean Genome and Epidemiology Study (KoGES), and Biobank Japan (BBJ). For African ancestry (AFR), we used data from the Uganda Genome Resource, MVP, and AoU (Supplementary Table 1).

In addition, we included the GWAS summary statistics for individuals of Admixed American (AMR), South Asian (SAS), and Middle Eastern (MID) ancestries. As large-scale GWAS data for these populations were limited, we conducted GWAS for BMI, WC, HIP, and WHR for these ancestry groups using the UK Biobank data. These analyses were performed using PLINK v2.0^70^, adjusting for sex, age, and the first ten principal components (PC) to control population stratification.

To increase the statistical power and enhance the resolution of association signals, we performed fixed-effects meta-analyses using METAL software^71^ for each anthropometric trait, when there are multiple GWAS summary statistics from different cohorts. Summary statistics from multiple sources were harmonised and combined following established quality control and meta-analysis protocols. The resulting summary statistics were used for subsequent genetic and functional analyses. A comprehensive list of the datasets and ancestry-specific details is provided in Supplementary Table 1.

### Multivariate GWAS of obesity using genomic SEM

To investigate the genetic architecture underlying obesity, we performed multivariate GWAS using genomic structural equation modelling (SEM)^13^. Summary statistics from GWAS of four anthropometric traits—BMI, WC, HIP, and WHR—were first harmonised and analysed within a hierarchical factor model. For each ancestry group (EUR, EAS, AFR, AMR, SAS, and MID), we performed separate multivariate GWAS using genomic SEM. SNP associations with the latent obesity factor were estimated using the diagonally weighted least squares (DWLS) fit function. As part of quality control, SNPs with minor allele frequency (MAF) ≤ 1% were excluded (Supplementary Tables 5 and 6). To ensure compatibility across datasets, only SNPs present in all ancestry-specific GWAS summary statistics and the corresponding ancestry-specific reference panels from the 1000 Genomes Project Phase 3 were retained^72^. Specifically, the EUR, EAS, AFR, AMR, and SAS ancestries were matched to their respective 1000 Genomes reference panels, whereas the MID ancestry was matched to a Middle Eastern reference panel derived from the UK Biobank. Finally, ancestry-specific multivariate GWAS summary statistics were meta-analysed using METAL^71^ to generate a comprehensive, large-scale, multi-ancestry GWAS for obesity.

### SNP-based heritability and genetic correlation estimation

We utilised LDSC v1.0.1 to estimate SNP-based heritability and bivariate genetic correlations among obesity-related traits—including BMI, WC, HIP and WHR—within each ancestry group. For each analysis, we used ancestry-specific precomputed LD scores derived from appropriate reference panels corresponding to each population. Approximately 1.3 million high-quality SNPs defined by the HapMap3 Consortium were included in the analyses. Genomic inflation was evaluated using the LDSC intercept and attenuation ratio, with values close to 1 and 0, respectively, suggesting that inflation in the test statistics was largely attributable to polygenicity rather than to confounding factors due to population stratification. Bivariate genetic correlations were calculated among the BMI, WC, WHR, and HIP GWAS results derived using genomic SEM (Supplementary Table 4).

### Defining obesity-associated signals and loci

To define the independent genetic signals associated with obesity, we identified all single-nucleotide variants (SNVs) that reached genome-wide significance (P < 5 × 10^-8^) in a multi-ancestry, multi-trait GWAS. Lead loci were defined by first identifying index SNVs using a greedy clumping algorithm implemented in PLINK v2.0, ranking SNVs by ascending P value. Clumps were constructed by assigning SNVs within 5 Mb of an index SNV, if they exhibited linkage disequilibrium (LD; *r*^2^ > 0.05) with the index SNV in at least one of the five continental reference populations from phase 3 of the 1000 Genomes Project.

Index SNVs located within 1 Mb of each other were grouped into single loci. Each locus was defined as a genomic region extending 500 kb upstream and downstream of the outermost index SNVs within that locus. A locus was considered previously reported if it overlapped with regions identified at genome-wide significance in earlier large-scale GWASs of obesity-related traits (Supplementary Table 2).

### Effector gene prioritisation using FLAMES

To identify candidate effector genes underlying the genetic loci associated with obesity across multiple ancestries, we applied the FLAMES (Fine-mapped Locus Assessment Model of Effector Genes) framework, as described by Schipper et al.^16^. FLAMES integrates SNP-to-gene annotations and convergence-based evidence to assign a probabilistic score to genes within each fine-mapped locus, reflecting their likelihood of mediating trait-associated genetic variations.

We first performed fine mapping of the loci identified in our multi-ancestry, multi-trait GWAS on obesity, defining credible sets of putative causal variants for each locus. These finely mapped loci served as inputs to FLAMES, which utilises a gradient-boosted machine learning model (XGBoost) trained on independent ExWAS-implicated gene-locus pairs. SNP-to-gene annotations, including chromatin interaction data, expression QTLs, and enhancer–promoter links, were integrated into the model alongside convergence-based prioritisation scores (PoPS).

Each gene in the locus was scored based on its functional relevance to the credible set and similarity to other trait-associated genes in genome-wide networks. Genes surpassing the recommended thresholds for both the scaled FLAMES score (>0.248) and raw FLAMES score (>0.136) were prioritised as candidate effector genes (Supplementary Table 3). This approach enables systematic and biologically informed prioritisation of genes that are most likely to mediate the observed obesity associations across diverse populations.

### Cross-ancestry comparison of effect sizes and MAF

To assess the consistency of GWAS signals across ancestries, we compared lead SNPs identified in EUR, EAS, and AFR using summary statistics from ancestry-specific multi-trait GWAS. For each ancestry (EAS and AFR), we selected genome-wide significant lead SNPs and extracted the corresponding effect sizes (beta) and SEs from the EUR results. For SNPs shared between EUR and each ancestry, we performed a linear regression of EUR beta on EAS or AFR beta and calculated R^2^to quantify the correlation. We also computed the 95% confidence interval (CIs) and visualised them using scatter plots and error bars. Additionally, we compared the MAFs between EUR and each ancestry. SNPs that were uniquely significant in non-EUR ancestries are highlighted separately. This approach allowed us to evaluate ancestry-specific and shared genetic signals across EUR, EAS, and AFR.

### Ancestry-specific clustering of obesity-associated genetic variants

For our obesity-focused analyses, we performed a cluster analysis using GWAS summary statistics generated via our multi-ancestry, multi-trait GWAS. For the multi-ancestry clusters, GWAS-significant (P < 5 × 10^-8^) variants were extracted from the multi-ancestry, multi-trait GWAS. Cluster analyses were also conducted separately for each EUR, EAS, and AFR ancestry group. The same significance threshold was applied to extract variants for the ancestry-specific cluster analyses based on the EUR, EAS, and AFR multi-trait GWAS results. Although the multi-ancestry clustering incorporated individuals of the EUR, EAS, AFR, AMR, SAS, and MID ancestries, ancestry-specific clustering was not performed for the AMR, SAS, or MID groups because of the absence of genome-wide significant variants in their respective multi-trait GWAS.

After excluding indels and variants located in the major histocompatibility complex (MHC) region, the remaining variants were subjected to five independent rounds of LD pruning (LD *r*^2^ < 0.05, MAF < 0.001), one for each ancestry-specific reference panel. Only variants that were identified as independent across all five populations were retained. Variants with high missingness across GWASs, multi-allelic variants, or ambiguous alleles (A/T and C/G) were replaced with high-LD (*r*^2^ > 0.8) proxies. As a final filter, variants were cross-referenced with the largest multi-ancestry obesity GWAS and removed if they had P > 0.05 or showed discrepancies in reported risk alleles. The final set of 1092 obesity-associated variants is shown in Supplementary Table 10.

To construct a matrix suitable for genetic clustering, we initially compiled GWAS for 75 obesity-related continuous phenotypes (lipid levels, diabetic traits, etc) (Supplementary Table 9). For each obesity-related trait, we prioritised multi-ancestry meta-analyses when available and otherwise defaulted to European ancestry GWAS (Supplementary Table 9). The traits exhibiting a high pairwise correlation (Pearson’s r > 0.80) were excluded to minimise redundancy. When correlated traits were identified, we retained the trait that demonstrated the strongest variant–trait association signal (Supplementary Table 10c). This filtering process resulted in a final dataset comprising 1,092 genetic variants across 63 distinct obesity-related traits. Variant–trait associations were represented as standardised z-scores and oriented according to the allele associated with increased obesity risk. For variants lacking association statistics for a given trait, z-scores were imputed using proxy variants in linkage disequilibrium (LD *r*^2^ > 0.5), where available; otherwise, the median z-score for that trait was assigned.

### Biobank analyses

We performed individual-level analyses for partitioned polygenic scores (pPSs) using two large-scale biobank resources: the UK Biobank and the Mass General Brigham (MGB) Biobank. Importantly, both datasets are independent of the GWAS cohorts used to generate the clustering of variants, ensuring an unbiased evaluation of pPS associations.

In this study, the UK Biobank served as the primary discovery cohort, whereas the MGB Biobank was used for the external validation of the findings. In both cohorts, we computed cluster-specific pPSs by applying variant weights derived from cluster loadings—obtained via Bayesian nonnegative matrix factorisation (bNMF)—to individual-level genotypes. For multi-ancestry clusters, we applied a threshold of 0.717 to the variant weights to retain only those that contributed most strongly to the cluster-specific signal in accordance with approaches previously adopted to enhance the signal-to-noise ratio^3,37^.

In the UK Biobank, we analysed a subset of participants of European ancestry with available genotyping and relevant phenotypic information. We computed standardised values for quantitative traits, including BMI, WC, systolic and diastolic blood pressure (SBP, DBP), lipid profiles [high-density lipoprotein (HDL), low-density lipoprotein (LDL), triglycerides (TG)], liver enzymes [alanine transaminase (ALT), gamma-glutamyl transferase (GGT)], Kidney function was assessed by estimating the glomerular filtration rate (eGFR), which was calculated using the Chronic Kidney Disease Epidemiology Collaboration (CKD-EPI) equation^73^, and glycaemic biomarkers [glycated haemoglobin (HbA1c)]. Trait standardisation was performed using z-transformation. Binary outcomes, including breast cancer (BC), colorectal cancer (CC), coronary artery disease (CAD), heart failure (HF), hypertension (HYP), non-alcoholic fatty liver disease (NAFLD), and type 2 diabetes (T2D), were defined based on a combination of International Classification of Diseases, 10th Revision (ICD-10) codes and self-reported data. (Supplementary Table 14).

We then tested the associations between the standardised pPSs and a set of diseases and quantitative traits using linear and logistic regression models. Each model was adjusted for age, sex, and the first ten PCs. The discovery in UK Biobank and validation in MGB Biobank allowed us to assess the reproducibility and robustness of the associations across independent populations and healthcare systems.

To calculate the GWAS-partitioned polygenic score, we first extracted genetic variants from each cluster derived from earlier steps. For each cluster, we identified corresponding variants in the summary statistics of external GWASs for specific MRI traits and harmonized their alleles, as demonstrated elsewhere^3^. We then performed an inverse-variance weighted fixed-effects meta-analysis, weighting each variant by the inverse of its squared standard error, to estimate the cluster-level polygenic effects. This approach allowed us to quantify the aggregate genetic effect of obesity-associated loci on a set of imaging-derived phenotypes, including gluteofemoral adipose tissue (GFAT), visceral adipose tissue (VAT), abdominal subcutaneous adipose tissue (ASAT), epicardial adipose tissue (EPAT), liver fat percentage, pancreatic fat percentage, traits that reflect fat distribution, and ectopic fat accumulation, which are closely associated with obesity-related metabolic risk.

All analyses were performed in accordance with the relevant ethical regulations, and institutional review board (IRB) approval was obtained from both the UK Biobank and MGB Biobank. All the participants provided informed consent for the use of their genomic and clinical data.

### Comparative analysis of genetically enriched obesity endotypes in overweight and obese populations

To investigate the phenotypic differences across genetically defined obesity endotyped groups, we utilised pre-computed pPS. The Hypothalamic dysregulation cluster was used as the reference group, as it most closely reflects features of conventional obesity. To emulate the clinical scenario where individuals with overweight or obesity present for care, we confined the analysis to UK Biobank participants with a BMI > 25 kg/m^2^. To assess the distinct clinical and metabolic characteristics associated with each endotype, we conducted logistic and linear regression models, comparing each outcome for endotyped group (top 20% of each endotype) versus control group (top 20% of Hypothalamic dysregulation cluster) (Extended Data Figs. 4 and 5). A range of phenotypic traits were assessed, including anthropometric, metabolic, hepatic, and renal measures, such as BMI, WC, LDL, HDL, TG, SBP, DBP, ALT, GGT, HbA1c, and eGFR. Continuous variables were standardised as z-scores, and for some analyses, they were transformed into decile-based ordinal variables. All models were adjusted for age and the first ten principal genetic components. To assess potential sex-specific effects, analyses were conducted in the overall cohort and separately in the male and female subgroups.

### Enrichment of obesity-associated SNPs for cell-type-specific open chromatin within clusters

We performed cluster-specific enrichment analyses using a matched set of null SNPs to assess cluster-associated genetic signals enrichment for cell type–specific regions of open chromatin. For each cluster of obesity-associated index SNPs (hereafter, “lead SNPs”), we defined null SNPs as variants located within 50 kb of the lead SNP that were not in LD with any lead SNP in the cluster (defined as *r*^2^ < 0.05) based on the 1000 Genomes Project phase 3 reference data. This strategy was applied independently to each cluster to maintain the specificity of its genetic architecture. The Hypo LpA and Proinsulin clusters were excluded from the cluster-specific enrichment analysis because of an insufficient number of variants, which made it infeasible to construct a well-matched null SNP set. As enrichment testing relies on statistically robust comparisons to null variants matched on characteristics, such as minor allele frequency and linkage disequilibrium, the small variant count in these clusters rendered the analysis underpowered and methodologically unreliable.

We define a binary response variable *Y_j_*, which takes the value of 1 if the *j*th SNP is a lead SNP and 0 if it is a null SNP. SNPs were annotated based on their overlap with gene elements obtained from Ensembl release 104^74^, including protein-coding exons, five untranslated regions (UTRs), and three UTRs. For each SNP, we defined the indicator variables G*_j_*^EXON^, G*_j_*^5UTR^, and G*_j_*^3UTR^, which take the value 1 if the SNP is mapped to the respective genic annotation and 0 otherwise. We further mapped the SNPs to regions of accessible chromatin using single-cell ATAC-seq peak sets from the CATLAS database, which comprises 222 cell types across 30 adult and 15 fetal human tissues, as well as 106 cell types from the adult human brain^75,76^. For each cell type *i*, we defined a binary indicator variable *X_ij_*, which takes the value of 1 if SNP *j* overlaps an ATAC-seq peak in cell type *i*, and 0 otherwise. Within each cluster, we modelled the enrichment of lead SNPs for cell type–specific ATAC-seq peaks using Firth bias-reduced logistic regression, adjusting for genetic annotations. The model is specified as follows:

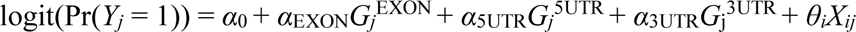

Here, α_0_ is the intercept, α_EXON_, α_5UTR_ and α_3UTR_ represent log-odds enrichment for genic annotations, and *θ_i_* captures the log-odds enrichment for overlap with cell type *i*’s accessible chromatin. We tested the significance of enrichment in each cell type by comparing the deviance of the full model (with unconstrained *θ_i_*) to a reduced model with *θ_i_* = 0. Significance thresholds were adjusted using the Bonferroni correction for the number of cell types tested. All enrichment models were implemented using the Logistf R package.

### Gene expression profiling across cell clusters

To characterise the cell type-specific expression patterns of enriched genes in each cluster, we analysed processed single-cell RNA-sequencing (scRNA-seq) data from six human tissues: adipose tissue^77^, brain^78^, heart^79^, liver^80^, pancreas^81^, and skeletal muscle^82^. All datasets were preprocessed and integrated using the Seurat framework (v4.3.0)^8^, and cell identities were annotated based on curated cell type labels from the original publications. For each tissue, gene expression data were extracted from the RNA assay of the corresponding Seurat object. We focused on 11 clusters derived from multi-ancestry, multi-trait obesity GWAS. For each cluster, we evaluated expression profiles across all annotated cell types (i.e., Seurat clusters) by calculating two metrics: (i) the mean log-transformed average expression of cluster genes [log₂(1 + average expression)], and (ii) the mean proportion of cells expressing each gene within the cluster (defined as the proportion of cells with a raw count ≥ 1), averaged across all genes. Expression values were summarised using the AverageExpression function implemented in Seurat (Supplementary Table 26).^3^

### MAGMA tissue expression analysis

Tissue-specific gene expression analyses were performed using the MAGMA^84^ tool in FUMA^85^. The analysis was conducted across 53 tissue types based on GTEx v8 data^86^. MAGMA was used to test whether obesity-associated genes were more highly expressed in specific tissues than expected, thereby providing insights into tissue-specific genetic regulation.

### Cell- and tissue-type-specific heritability enrichment analysis

We performed cell- and tissue-type-specific heritability enrichment analyses using LDSC-SEG^87^. The analysis was restricted to tissue-specific gene expression profiles from the GTEx v8 data. The LDSC-SEG evaluates whether the heritability of a trait is enriched in genes with high tissue-specific expression. The human leukocyte antigen (HLA) region was excluded from all analyses due to its complex linkage disequilibrium structure. We utilised precomputed LD score annotation files, as described by Finucane et al.^87^, which are publicly available and specific to the GTEx v8 dataset.

### Pathway enrichment analysis of obesity-associated genes identified from multi-ancestry, multi-trait GWAS

Pathway and functional enrichment analyses were performed for genes associated with obesity in the multi-ancestry, multi-trait GWAS using gprofiler2 v.0.2.3^88^. The analysis included Gene Ontology (GO) terms—biological process, molecular function, and cellular components—as well as additional functional annotation databases, including Human Phenotype Ontology (HP), Human Protein Atlas (HPA), Kyoto Encyclopedia of Genes and Genomes (KEGG), transcription factor targets (TF), and WikiPathways (WP). A two-stage enrichment algorithm implemented in gprofiler2 was used to identify the significantly overrepresented terms and pathways. Statistical significance was assessed using a cumulative hypergeometric test, and P-values were adjusted for multiple testing using Bonferroni correction. Enriched terms were interpreted to highlight the potential biological mechanisms underlying genetic susceptibility to obesity.

## Data availability

The GWAS summary statistics for this study are available at https://www.ebi.ac.uk/gwas. The PRS weights developed for the multi-ancestry obesity GWAS are available from the PGS Catalog (https://www.pgscatalog.org/). All referenced GWAS summary statistics are publicly available and cited in Supplementary Tables 1 and 9. Individual-level phenotypic and genetic data for the UKB (https://www.ukbiobank.ac) and MGB (https://www.massgeneralbrigham.org/en/research-and-innovation/participate-in-research/biobank) can be accessed through this application. The reference panel for 1kGp3 was obtained from https://mathgen.stats.ox.ac.uk/impute/1000GP_Phase3.html. The epigenomic activity databases are available online for CATLAS (https://catlas.org).

## Code availability

All software used in the study is publicly available as described in the “Methods.” Analyses were conducted using the following publicly available software: METAL v.2011-03-25 (https://genome.sph.umich.edu/w./METAL), PLINK v.2.0 (https://www.cog-genomics.org/plink/2.0/), genomic SEM software (https://github.com/GenomicSEM/GenomicSEM), and LDSC software, including S-LDSC and SEG-LDSC (https://github.com/bulik/ldsc). The LDScore is available at https://alkesgroup.broadinstitute.org/LDSCORE and FUMA at https://fuma.ctglab.nl. The code for variant preprocessing, bNMF clustering, and basic visualisation is available at https://github.com/gwas-partitioning/bnmf-clustering.

## Acknowledgments

S.J.C. was supported by a grant from the National Heart, Lung, and Blood Institute (K99HL177340). P.N. was supported by grants from the National Institutes of Health (R01HL127564 and U01HG011719) and the National Heart Lung and Blood Institute (K08HL161448 and R01HL164629). P.T.E. was supported by grants from the National Institutes of Health (1R01HL092577, 1R01HL157635, and 5R01HL139731), the American Heart Association Strategically Focused Research Networks (18SFRN34110082), and the European Union (MAESTRIA 965286). The A.C.F. was supported by grants from the National Heart, Lung, and Blood Institute (K08HL161448 and R01HL164629). We gratefully acknowledge all participants of the UKB and MGB Biobanks for their contributions, without whom this research would not have been possible.

## Author contributions

Concept and design: M.S.K., M.S., and H.W. Acquisition, analysis or interpretation of data: M.S., M.S.K., H.K., S.P., I.S, S.L., B.K., X.C., Y.S., P.T.E., A.C.F., and H.W. Statistical analysis: M.S. and M.S.K. Drafting of the manuscript: M.S.K. and M.S. Critical revision of the manuscript for important content: M.S.K., M.S., S.M.J.C., S.K., P.N., P.T.E., A.C.F., and H.W. Administrative, technical, or material support: P.T.E, A.C.F., and H.W. Resources: P.T.E., A.C.F. and H.W. Supervision: P.T.E, A.C.F., and H.W.

## Competing interests

P.T.E. has received sponsored research support from Bayer AG, IBM Health, Bristol Myers Squibb, Pfizer, and Novo Nordisk. P.T.E. also served on advisory boards and consulted Bayer AG, MyoKardia, and Novartis. P.N. reports research grants from Allelica, Amgen, Apple, Boston Scientific, Cleerly, Genentech / Roche, Ionis, Novartis, and Silence Therapeutics, personal fees from Allelica, Apple, AstraZeneca, Bain Capital, Blackstone Life Sciences, Bristol Myers Squibb, Creative Education Concepts, CRISPR Therapeutics, Eli Lilly & Co, Esperion Therapeutics, Foresite Capital, Foresite Labs, Genentech / Roche, GV, HeartFlow, Magnet Biomedicine, Merck, Novartis, Novo Nordisk, TenSixteen Bio, and Tourmaline Bio, equity in Bolt, Candela, Mercury, MyOme, Parameter Health, Preciseli, and TenSixteen Bio, and spousal employment at Vertex Pharmaceuticals, all unrelated to the present work.. A.C.F. reports being the co-founder of Goodpath, serving as scientific advisor to MyOme and HeartFlow, and receiving a research grant from Foresite Labs. The authors declare no conflicts of interest.

---

**Extended Data Fig. 1.**
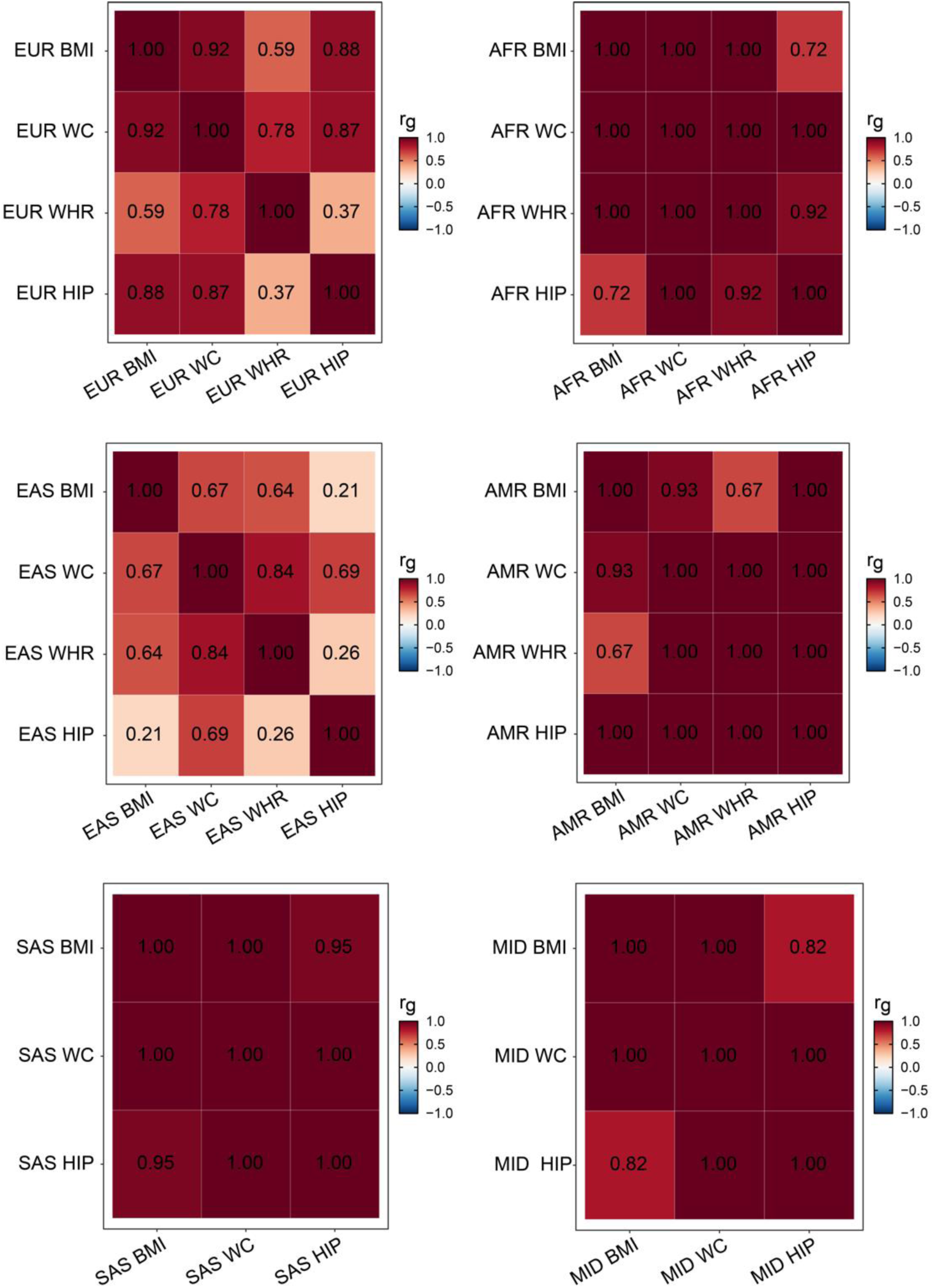
Genetic correlation of obesity traits across diverse ancestries in a multivariate genetic factor model. Genetic correlations among the obesity components included in the multivariate genetic factor model across different ancestries were estimated using LD score regression, with the colour bar representing the genetic correlation values. The diagonal indicates SNP-based heritability, and the upper and lower off-diagonal triangles represent pairwise genetic correlations and standard errors, respectively.

**Extended Data Fig. 2.**
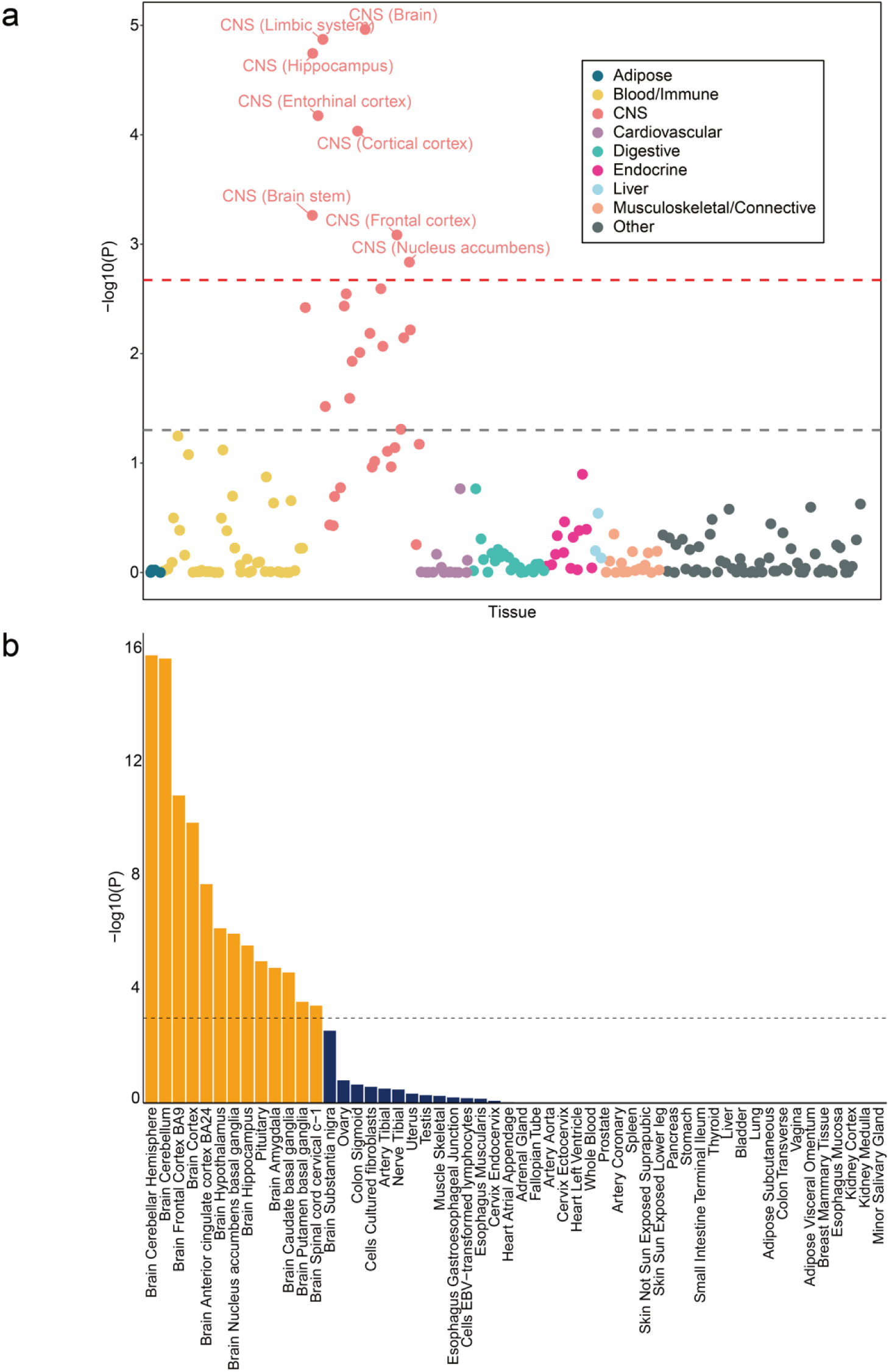
Tissue-specific enrichment analysis results for multi-ancestry obesity. **a** Tissue-specific enrichment analysis across multiple tissues based on gene expression using stratified linkage disequilibrium score regression (LDSC-SEG). Each dot represents gene expression data from a specific tissue or cell type sourced from either the GTEx or Franke lab, with colours indicating the tissue categories. The x-axis represents tissue categories, whereas the y-axis shows the uncorrected log_10_(*P*) values from one-sided z-tests for enrichment. The red dashed line at –log_10_(*P*) = 2.3 marks the false discovery rate (FDR) threshold of 5%, indicating statistically significant tissue enrichment, while the black dashed line at –log_10_(*P*) = 1.3 represents the nominal significance threshold of *P* < 0.05. Tissues surpassing the FDR threshold were labelled. Significance was determined using two-sided tests, and *P* values were adjusted using the Benjamini-Hochberg FDR correction. **b** Tissue-specific gene expression analysis using MAGMA. The bar plot illustrates the statistical significance of tissue enrichment of obesity-related genes based on GTEx v8 gene expression data. The x-axis represents different tissues, while the y-axis shows-log_10_(*P*) values from gene-set analysis (GSA). Bars are coloured based on statistical significance: tissues with –log_10_(*P*) > 2.55 are highlighted in orange, while others are shown in dark blue. A dashed horizontal line at –log_10_(*P*) = 2.55 denotes the threshold for statistical significance (P < 0.001). False discovery rate (FDR) correction was applied using the Benjamini-Hochberg method, with an FDR < 0.05, corresponding to *P* < 0.00279 (–log_10_(*P*) = 2.55). Statistical significance was determined by two-tailed tests.

**Extended Data Fig. 3.**
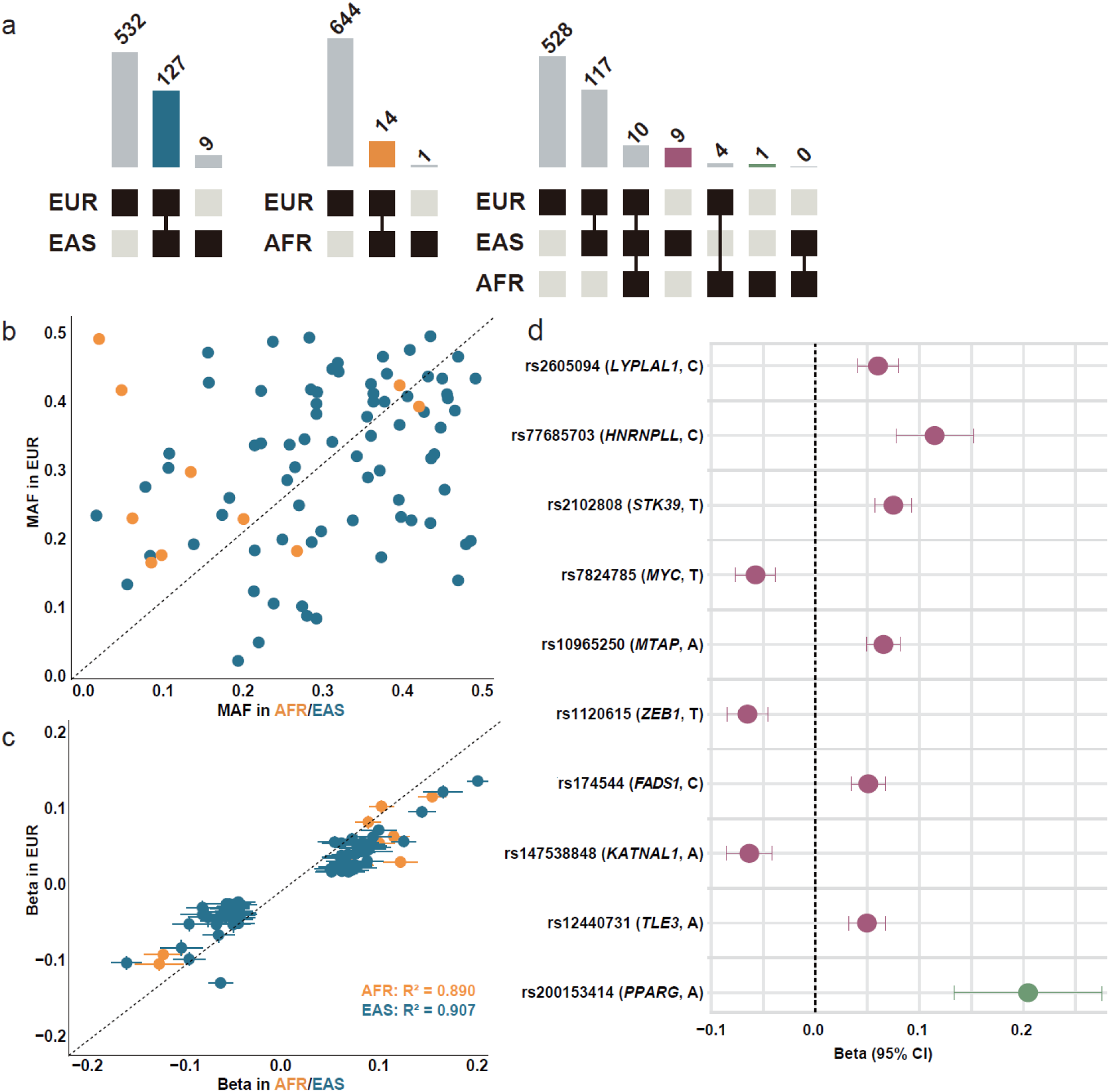
Shared and ancestry-specific allelic effects within loci across diverse populations. **a** The Upset plot illustrates the combinations of populations that detected obesity GWAS signals through intra-population meta-analysis. The bar chart at the top quantifies the number of GWAS loci across different population combinations, with each bar representing the total number of loci observed for specific combinations, as indicated by connected points in the central matrix. The central matrix displays the population combinations involved in each set of associations, where filled squares denote the populations included in a particular combination. The left panel compares the EUR and EAS, the middle panel compares the EUR and AFR, and the right panel presents a comparison of the EUR, EAS, and AFR. **b** and **c**, Each point represents a significant GWAS variant in both the non-EUR and EUR populations. Orange dots indicate significant variants in AFR and EUR, whereas dark blue dots represent significant variants in EAS and EUR. **b** Comparison of minor allele frequencies (MAF) across populations, with the vertical axis showing MAF in EUR and the horizontal axis showing MAF in AFR or EAS. **c** Comparison of effect sizes, with the horizontal axis representing EUR effect sizes and the vertical axis representing those in the AFR or EAS. Error bars denote 95% confidence intervals and *R*^2^ indicates the squared Pearson correlation coefficient. **d**. The forest plot illustrates the effect sizes of ancestry-specific GWAS-significant variants that were not observed in other populations. The vertical axis displays each variant’s rs number, corresponding V2G gene symbol, and effect allele. Purple dots indicate variants observed exclusively in the EAS, whereas green dots represent variants observed only in the AFR. Error bars denote 95% confidence intervals.

**Extended Data Fig. 4.**
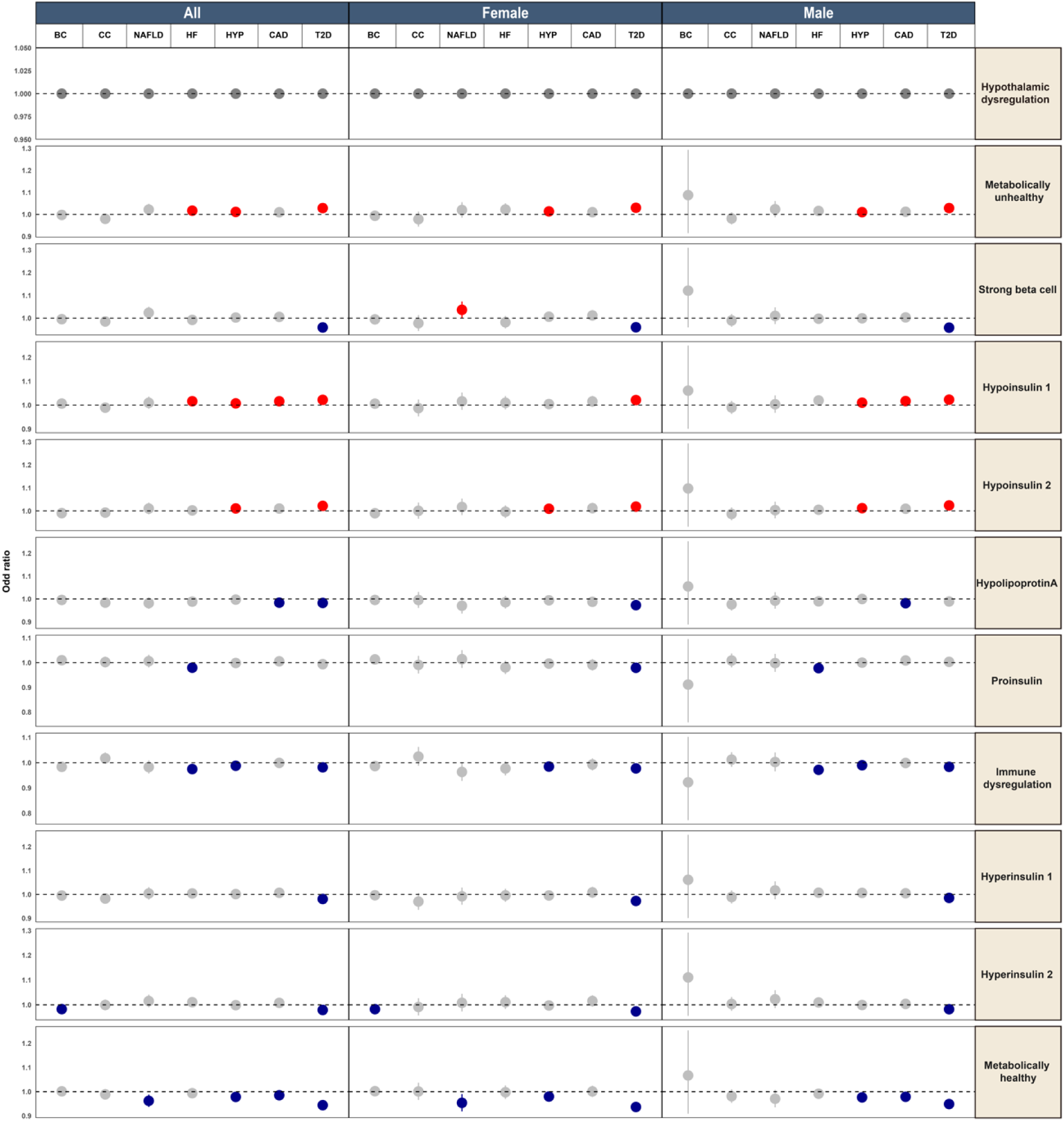
Forest plot of pPS associations with clinical health outcomes across obesity genetic clusters in overweight populations, stratified by sex. This figure illustrates the associations between pPS and various binary health outcomes in overweight populations. The analysis was stratified by sex (male, female) and the overall population (all), with logistic regression models estimating odds ratios (OR). Beta coefficients represent the log odds of each outcome associated with endotyped group (top 20% of each endotype) versus control group (top 20% of Hypothalamic dysregulation cluster). Each panel corresponds to a unique cluster (rows) and sex (columns), allowing a direct comparison of endotype-phenotype associations. The error bars depict 95% confidence intervals (CI). Colour coding was applied based on the statistical significance and direction of association: Red: Significant positive association (beta > 1, *p* < 0.05), indicating higher odds of disease; dark blue, significant negative association (beta < 1, *p* < 0.05), suggesting a protective effect; and Gray: Non-significant association (*p* ≥ 0.05). We set Hypothalamic dysregulation cluster as a reference, as it most closely reflects features of conventional obesity.

**Extended Data Fig. 5.**
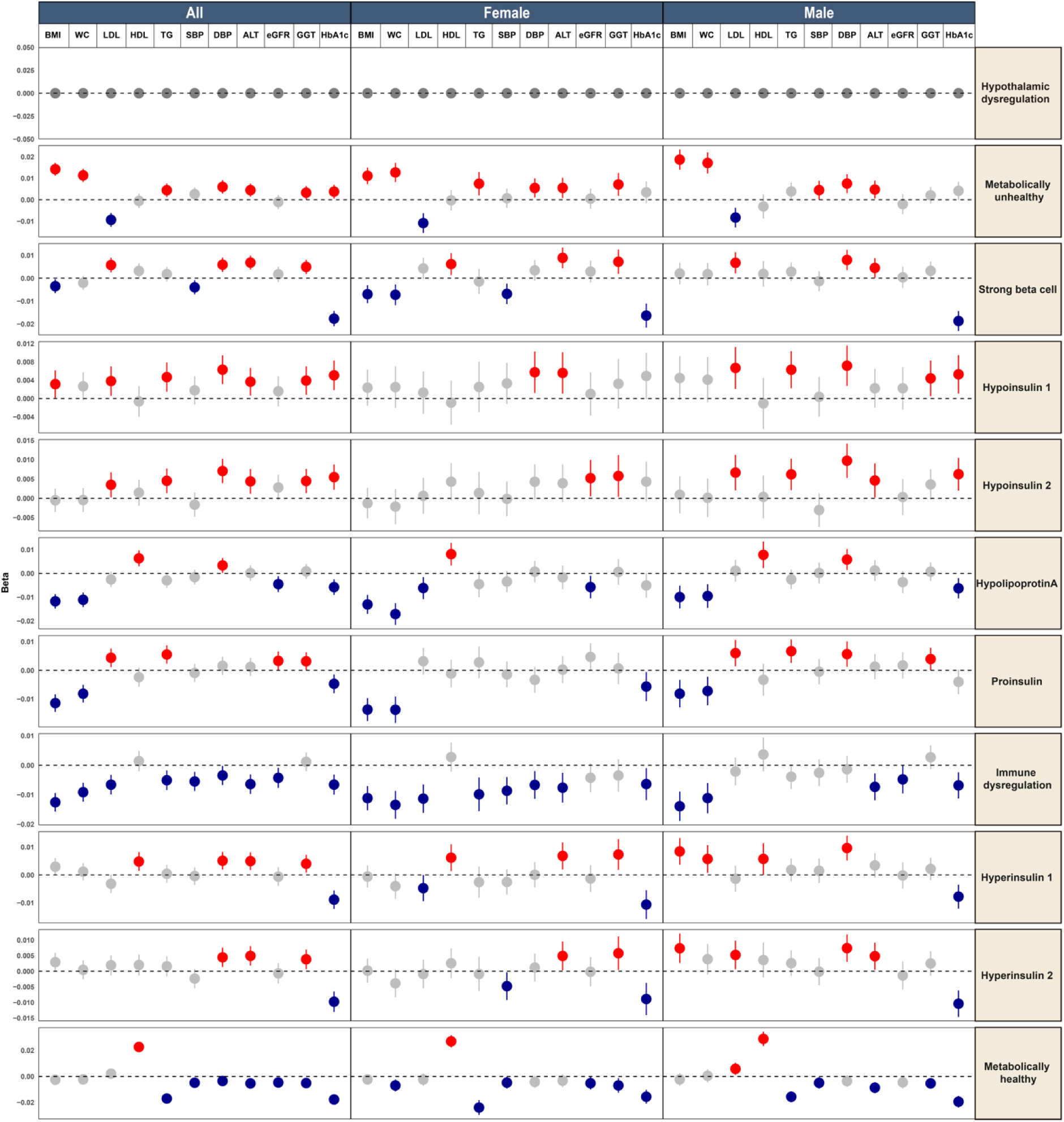
Forest plot of pPS associations with continuous metabolic traits across obesity genetic clusters in overweight populations, stratified by sex. This figure illustrates the associations between pPS and various continuous metabolic traits in overweight populations. The analysis was stratified by sex (male, female) and the overall population (all), with linear regression models estimating beta comparing endotyped group (top 20% of each endotype) versus control group (top 20% of Hypothalamic dysregulation cluster). Each panel corresponds to a unique cluster (rows) and sex (columns), allowing a direct comparison of endotype-phenotype associations. The error bars depict 95% confidence intervals (CI). Colour coding was applied based on the statistical significance and direction of association: Red: Significant positive association (beta > 1, *p* < 0.05), indicating higher odds of disease; dark blue, significant negative association (beta < 1, *p* < 0.05), suggesting a protective effect; and Gray: Non-significant association (*p* ≥ 0.05). We set Hypothalamic dysregulation cluster as a reference, as it most closely reflects features of conventional obesity.

**Extended Data Fig. 6.**
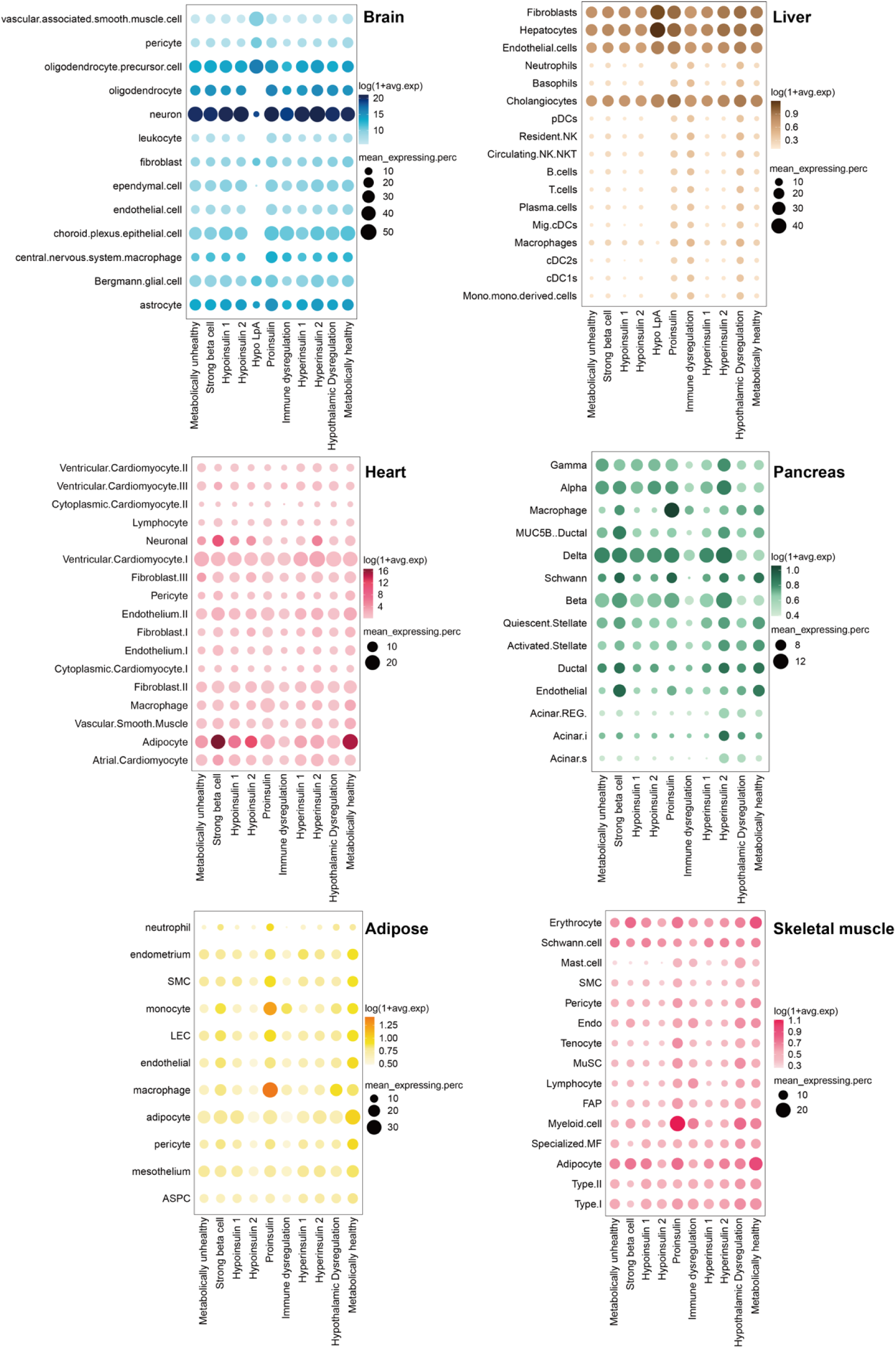
Integration of multi-ancestry obesity genetic clusters and transcriptomics with cell-type-specific analysis reveals distinct cell-type expression patterns for each cluster. Dot plot illustrating the mean expression of genes from obesity genetic cluster-specific components across six tissues (brain, liver, heart, pancreas, adipose tissue, and skeletal muscle) using single-nucleus transcriptomics. Each point represents a cluster-specific gene set, where the x-axis denotes cluster identity and the y-axis represents the different cell types. The colour gradient indicates *log(1+avg.exp)*, which represents the log-transformed average expression level of the genes in each cluster. The size of each point corresponds to *mean_expressing.perc*, which is the average percentage of cells within a cluster that express genes from the given gene set. Larger points indicate a higher proportion of cells expressing these genes, whereas darker colours represent higher average expression levels.

**Extended Data Fig. 7.**
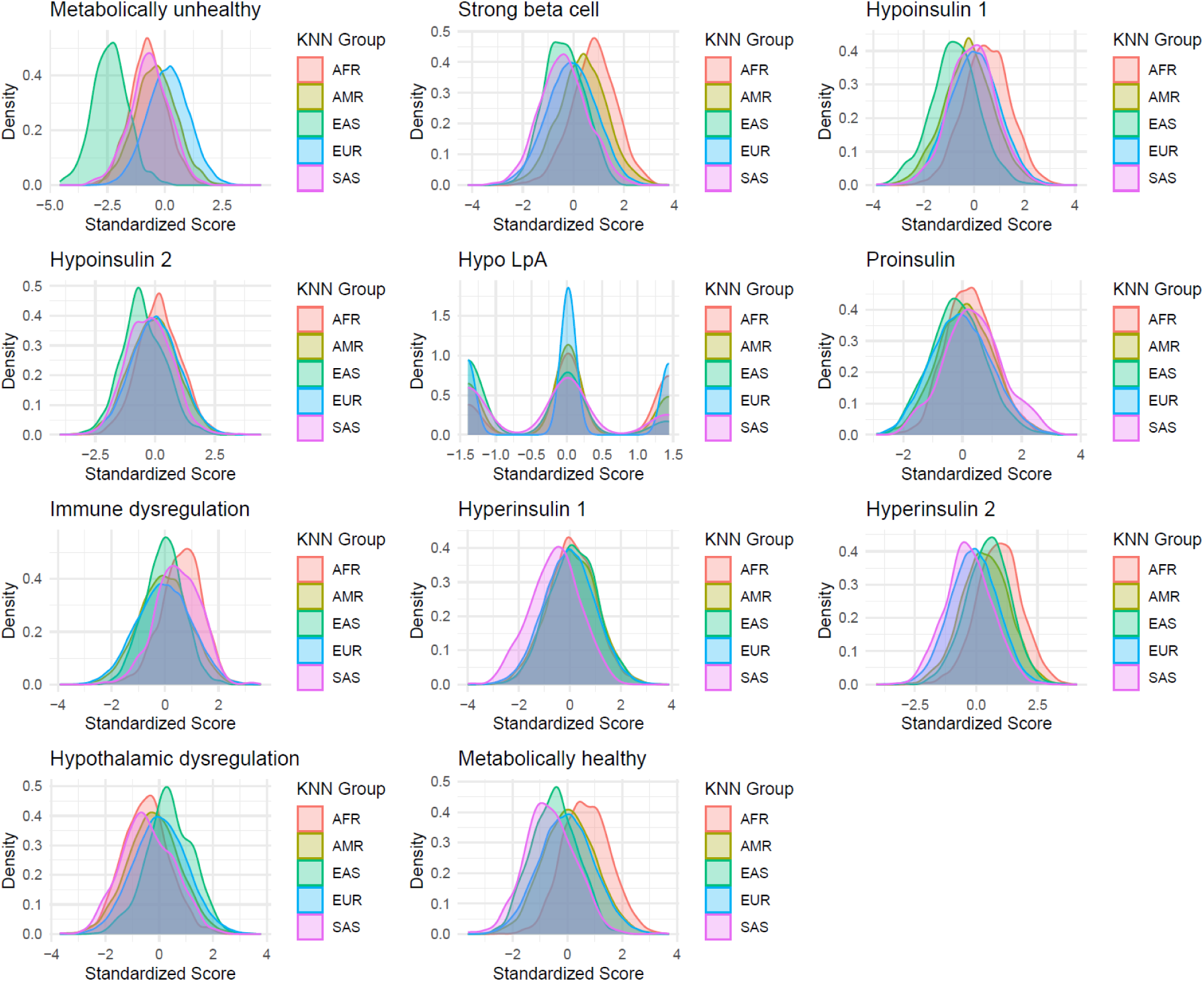
Variation in distribution of multi-ancestry obesity genetic clusters across ancestry groups before adjustment for principal components. Each histogram displays the pPS distribution for the indicated multi-ancestral obesity genetic clusters. For each cluster, the pPS for the entire cohort was standardised to a normal distribution and a separate curve was displayed for each genetically inferred ancestry group. All analyses were performed using MGB Biobank. AFR, African; AMR, Admixed American; EAS, East Asian; EUR, European; SAS, South Asian.

**Extended Data Fig. 8.**
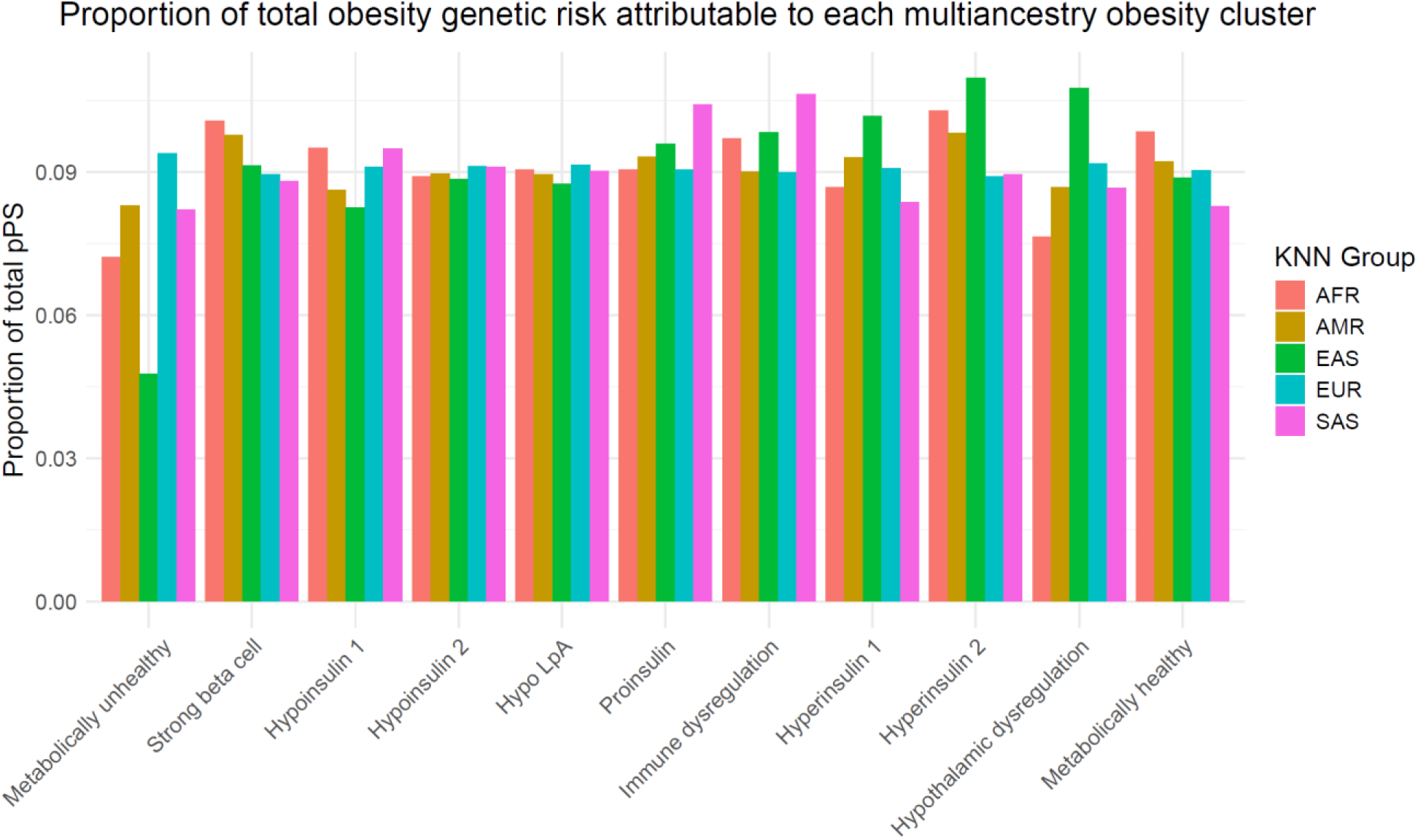
Proportion of total obesity genetic risk attributable to each multi-ancestry obesity cluster in MGB Biobank. For each individual, the total obesity genetic risk was calculated as the sum of the pPS across the 11 multi-ancestral obesity genetic clusters. All the individuals were grouped according to their genetically inferred ancestry. Each bar displays the proportion of total obesity genetic risk conferred by each cluster. AFR, African; AMR, Admixed American; EAS, East Asian; EUR, European; SAS, South Asian.

**Extended Data Fig. 9.**
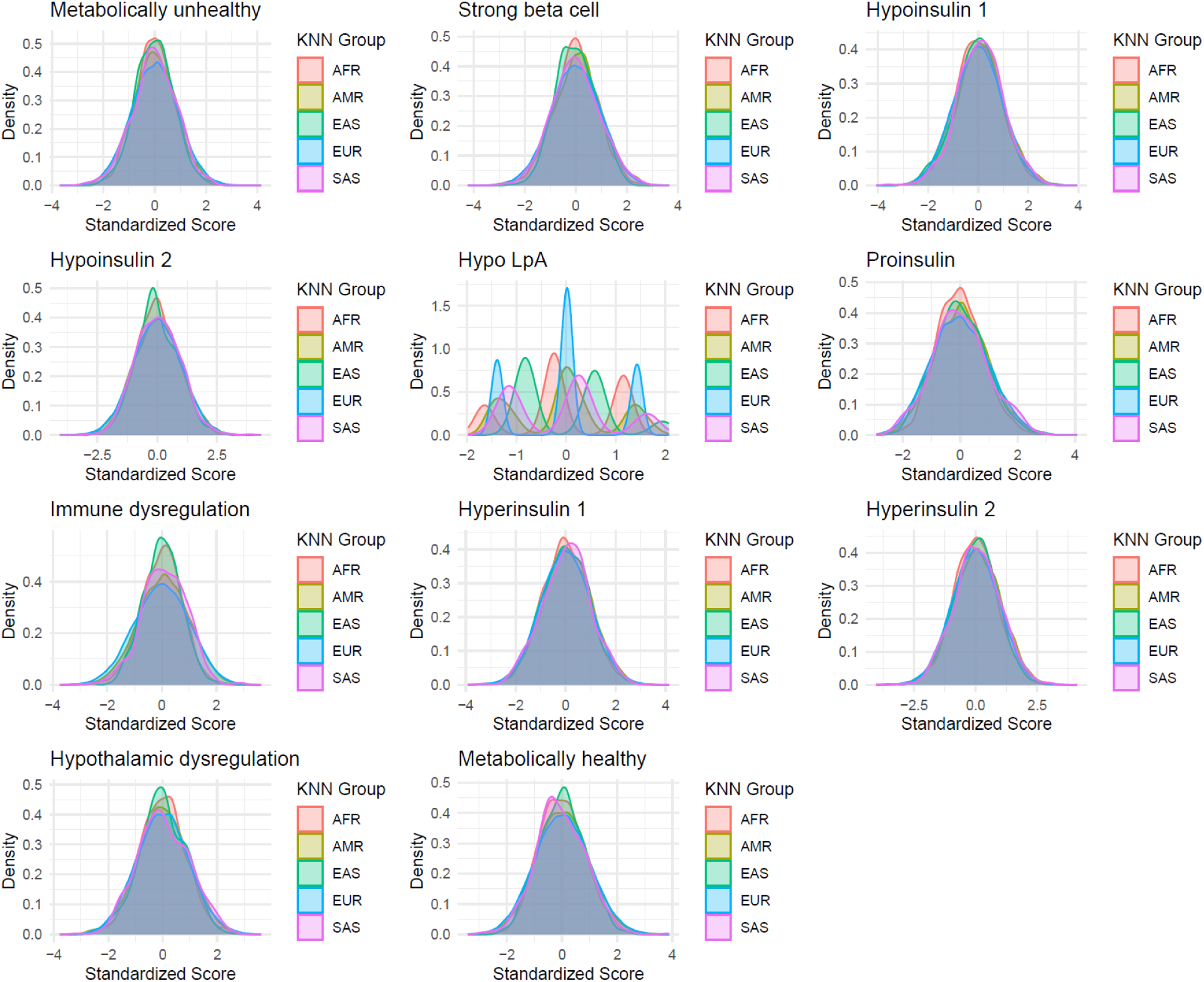
Distribution of multi-ancestry obesity genetic clusters across ancestry groups after adjustment for principal components (PC). Each histogram displays the pPS distribution for the indicated multi-ancestral obesity genetic clusters. For each cluster, the top 10 PCs were regressed from the pPS and standardised to a normal distribution. We used PC-adjusted pPS in any analysis that required ancestry adjustment for the direct comparison of clustered individuals. A separate curve is displayed for each genetically inferred ancestry group. All analyses were performed using MGB Biobank. AFR, African; AMR, Admixed American; EAS, East Asian; EUR, European; SAS, South Asian.

## Notes

### Competing Interest Statement

The authors have declared no competing interest.

### Summary of Updates

We have updated author list

